# Prediction of Individual Speech Outcomes with Cochlear Implants: Limitations and Opportunities for Clinical Translation

**DOI:** 10.64898/2026.09.07.26362458

**Authors:** Marta Campi, Alexander Huber, Tobias Goehring

## Abstract

**Objectives:** Speech outcomes with cochlear implants (CIs) vary widely across patients and remain difficult to predict from pre-operative data. This study addressed three questions with clinical relevance: (RQ1) Can AI models predict individual CI speech outcomes, and if not, why? (RQ2) Can pre-operative data be used to identify patients at risk of poor outcomes? (RQ3) Do post-operative speech scores early after implantation contain prognostic information for later speech outcomes?

**Design:** A retrospective analysis was performed using the Swiss National Cochlear Implant Database, Zurich centre (*N* = 771 patients; 473 adults after exclusion of single-sided deafness). For RQ1, pre-operative data and seven prediction methods, from standard linear regression to recent machine-learning models, were used to predict post-operative word recognition scores. Prediction models were assessed on held-out data and on data from patients implanted later than the cohort used for model training. For RQ2, the best-performing model under RQ1 was used to predict patients with a poor outcome after implantation (word-recognition score below 40%), and pre-operative variables were grouped by clinical type to identify sources of information for risk prediction. For RQ3, the prognostic value of early post-operative speech measurement was assessed by adding six-month post-operative word scores to the pre-operative data and a clustering analysis was performed to identify distinct trajectories post implantation.

**Results:** Cross-validated *R*^2^ for individual CI speech score prediction was small across all algorithms (best model with full feature set: *R*^2^ = 0.22) and conformal prediction intervals spanned ±41 percentage points, covering nearly the whole range of possible scores. Speech score distributions of patient groups defined by audiometric, aided-field and pre-operative speech variables converged after implantation (mean distance between group distributions decreased from 21.5 to 6.0 percentage points), while distributions of groups defined by developmental and communicative variables diverged. Pre-operative speech scores were much better predicted from pre-operative variables (*R*^2^ = 0.48 based on audiometric data only). For risk prediction, the best-performing model identified poor outcome with AUC = 0.71. Self-rated articulation was the dominant pre-operative predictor. Audiometric, aided-field, and pre-operative speech data on which other prediction models have relied classified poor outcome at chance (AUC = 0.53), whereas developmental and communicative variables performed better (AUC = 0.65). Adding the six-month post-operative word score to the pre-operative data substantially raised *R*^2^ for long-term outcome from 0.03 to 0.48. Four trajectory types (Early Responders, Late Responders, Non-Responders, and Decliners) emerged from the clustering analysis but were not predictable from pre-operative data.

**Conclusions:** The limited prediction performance for CI speech outcomes appears to be a property of the pre-operative data rather than of the prediction methods applied to them. However, pre-operative data can partially identify patients at risk of poor outcome. Importantly, speech data obtained within six months after implantation significantly improves outcome prediction at later time points. Longitudinal assessments post implantation therefore have potential for clinical intervention to improve long-term CI outcome. These findings support a two-stage framework for clinical translation: data-based risk counselling before surgery and post-operative monitoring for selective intervention after surgery.

## 1 Introduction

Cochlear implants are the most successful neural prosthesis developed to date, with over one million devices implanted worldwide (Zeng, 2022). For most recipients, the intervention provides access to spoken language that would otherwise be unavailable, with substantial improvements in speech perception, communication, and quality of life well documented (Boisvert et al., 2020). Over the past decades, many research efforts have been made to enhance CI performance further and to improve our understanding of the underlying limitations (Carlyon and Goehring, 2021). Yet individual outcomes vary enormously: post-operative speech scores for word recognition tests span the full range from 0 to 100%, and this variability persists even among patients with comparable clinical profiles (Lazard et al., 2012; Blamey et al., 2013). Roughly one in six adult recipients achieves poor outcomes (Demyanchuk et al., 2025), and individual performance remains largely unpredictable from pre-operative data, while post-operative factors such as electrode placement may better explain outcomes (Finley et al., 2008). As candidacy criteria expand to include patients with more residual hearing, older age, and shorter durations of deafness (Blamey et al., 2013), the clinical need has increased to counsel candidate’s expectations realistically and identify candidates at high risk of poor outcomes before surgery, as well as administer post-operative rehabilitation most effectively.

Pre-operative predictors of CI outcome have been studied extensively for over three decades. The factors most consistently associated with post-operative speech perception are duration of severe-to-profound hearing loss, age at implantation, etiology, and residual hearing, identified across numerous studies spanning multiple centres and continents (Lazard et al., 2012; Holden et al., 2013; Goudey et al., 2021). The landmark analysis by Blamey et al. (1996) reported that these factors explained 21% of the variance across 808 recipients, but when the same analysis was repeated seventeen years later with 2,251 recipients, the explained variance had dropped to approximately 10% (Blamey et al., 2013; Zhao et al., 2020; Stronks et al., 2025). The decline may reflect the expansion of candidacy criteria: as patients are implanted earlier and with more residual hearing, the variance that these predictors capture has been likely reduced further. Attempts to move beyond standard clinical variables have not resolved this, with neuroimaging and genetic markers producing high reported accuracies only in small, unvalidated studies (Nair et al., 2025; van de Velde et al., 2021). Despite three decades of effort, the proportion of variance explained by pre-operative factors has not substantially improved.

Machine learning methods have been increasingly applied to CI outcome prediction, motivated by their capacity to capture non-linear interactions that linear models cannot represent (Nair et al., 2025; Mo et al., 2025). However, the largest cross-validated studies show that algorithmic advancements do not yet overcome the limited prediction performance for CI outcomes. The most comprehensive analysis to date, covering 2,489 recipients across three clinics, found that gradient boosting reduced prediction error by less than one percentage point relative to linear regression, and that doubling the sample size improved accuracy by only 3% (Shafieibavani et al., 2021). Other large-scale studies confirmed this: cross-validated *R*^2^ stayed between 0.08 to 0.11 (Anthonisen et al., 2026), prediction error was around 17 to 22 percentage points regardless of algorithm choice (Shafieibavani et al., 2021; Demyanchuk et al., 2025), and machine learning methods achieved comparable performance to but did not exceed the predictive accuracy of experienced clinicians (Demyanchuk et al., 2025; Philpott et al., 2024). Interpretation of this literature is further complicated by the lack of standardisation across studies in outcome measures, evaluation metrics, and validation procedures (Mo et al., 2025; van de Velde et al., 2021). Nevertheless, there is a persistent limitation in prediction performance across previous studies. This raises a general question that motivated the present study: is it a limitation of the prediction algorithms, or a property of the data itself?

What could explain this limitation in prediction performance? The usual explanation is methodological: insufficient data overall, incomplete data, heterogeneous outcome measures, inadequate validation. These are real problems for the prediction of CI outcomes, but they are not sufficient to explain the occurrence and consistency of the finding. A prospective study by Peng et al. (2026) points toward a more fundamental issue: in a cohort of 43 new recipients (23 with complete measures), patient history variables explained none of the variance in speech outcomes (adjusted *R*^2^ = −0.054), while recipient-specific psychophysical and cortical measures (across-electrode intensity discrimination and cross-modal activation of the auditory cortex, both available only after implantation) explained 31%. This suggests that the variables available before surgery may not contain the information needed for prediction, not because they are poorly measured, but because cochlear implantation changes the relationship between pre-operative hearing status and post-operative speech perception: predictors that describe the pre-operative acoustic hearing system are unlikely to retain their predictive value once the cochlea is bypassed by direct electrical stimulation. To our knowledge this hypothesis has not been tested directly, nor has a structural account of the limitation in prediction performance for CI outcomes been proposed. If the limit is informational (ie. limited by the data) rather than methodological (ie. limited by the prediction model), then even the best possible models will face the same constraints and be limited in their performance. Calls to strengthen validation practices in this literature are well placed (van de Velde et al., 2021), but validation alone cannot recover information that the data do not contain. Previous work has largely interpreted poor predictive performance as a limitation of the algorithms or the study design. We show here that much of this limitation is informational: cochlear implantation selectively removes the predictive value of the variables on which the outcome prediction models rely most. We then ask whether pre-operative data *can* predict risk probability for poor outcomes rather than performance per se, reducing the task from a regression to a potentially simpler classification problem.

The same logic applies to the predicted outcome target and its clinical relevance. What counts as a cochlear implant “outcome” is in itself a complex question, with different measures being used across clinics and this lack of a shared prediction target is part of why results across studies are difficult to compare. We adopt monosyllable word recognition in quiet because it is what our dataset provides and what keeps our results comparable to the many cohorts discussed above. Therefore the limitations and analyses we report concern this outcome. With a view on clinical translation we investigate whether this outcome can be predicted as a continuous score for individual patients (Shafieibavani et al., 2021; Anthonisen et al., 2026; Stronks et al., 2025). As reported by previous studies this may fail and so we also follow a classification approach to test whether a patient at risk of a poor outcome can be identified before surgery, which is an important question during pre-operative counselling (Fraysse and James, 2019). If pre-operative information is not sufficient to guide and inform clinical decision-making reliably before CI implantation, we consider whether the missing information becomes available shortly after surgery and has predictive value to inform about and potentially help in improving a patient’s post-operative speech-perception trajectory with their CI.

In the present study, we used the Swiss National Cochlear Implant Database to address three research questions with clinical relevance. The first research question was whether pre-operative data can predict individual post-operative speech scores, and to analyse what causes the limited prediction performance. We implemented seven prediction algorithms with increasing capability, from linear regression to a foundation model (TabPFN (Hollmann et al., 2025)), and analysed how the link between pre-operative variables and CI outcome changes after implantation. We assessed whether cochlear implantation affects the predictive value of all pre-operative variables equally or selectively. The second research question was whether pre-operative data can identify patients at risk of a poor outcome: if continuous scores cannot be predicted well, binary risk may still be partially predictable, a coarser but clinically actionable target. The third research question was whether early post-operative speech scores contain prognostic information about long-term CI outcome. While pre-operative data may not be sufficient to forecast CI outcomes, early post-operative information could potentially improve predictions for long-term CI outcomes or guide rehabilitation for specific patient groups to improve their speech outcome trajectory.

## 2 Materials and Methods

### 2.1 Database and Participants

Data were drawn from the Swiss National Cochlear Implant Database (CICHDB), Zurich centre, which includes 771 patients implanted at University Hospital Zurich between May 2012 and November 2025 (Senn et al., 2018). The CICHDB is maintained as part of the Swiss highly specialised medicine (HSM) programme. Cochlear implantation in Switzerland is regulated tightly: under this programme only five designated centres are authorised to perform the procedure, and each is required by federal mandate to collect standardised demographic, audiometric, surgical, and speech test data for every implanted patient, with the data reported annually in anonymised form to the programme (Senn et al., 2018). The present study is restricted to adults (aged 18 years or older at implantation, *N* = 538). Patients with single-sided deafness (SSD), defined as a contralateral four-frequency pure-tone average (PTA; 500, 1000, 2000, 4000 Hz) of 30 dB HL or better, were further excluded (*N* = 65), yielding a study cohort of 473 adults with bilateral hearing loss. Of these 473 adults, the primary analyses used a subset of *N* = 378 with a post-operative monosyllable word-recognition score (WRS, the primary outcome measure; defined in Section 2.1.1) measured at least 6 months after implantation; a smaller subset of *N* = 264 adults with both pre- and post-operative WRS scores served as a sensitivity analysis (Section 2.2). The sample size for each analysis was determined by the completeness of data for that analysis. The sample sizes were 378 for RQ1 and RQ2, and 188 and 136 for RQ3; a paired subsample of 264 adults served as a sensitivity analysis. A complete map of the analyses, with the sample, predictors and outcome of each, is given in Supplementary Material, Table S1. Post-operative WRS was missing for 95 of the 473 adults. Availability of the outcome was not independent of the pre-operative profile: adults without a post-operative WRS score had poorer pre-operative hearing in the implanted ear, were less often native speakers of German or Swiss German, and more often had disturbed articulation, each independently of the others (Supplementary Material, Section S9). The implications are considered in Section 4. Audiometric thresholds that could not be measured (no response at the maximum output of the audiometer) are coded as 130 dB HL in the CICHDB. The maximum output is frequency-dependent and smaller at the low- and high-frequency ends of the audiogram, so 130 dB HL is a uniform coding convention rather than a measured level; the univariate screen of audiometric and aided-field variables therefore excludes these thresholds rather than imputing them (Supplementary Material, Table S3). Demographic and clinical characteristics are summarised in Table 1.

**Table 1:**
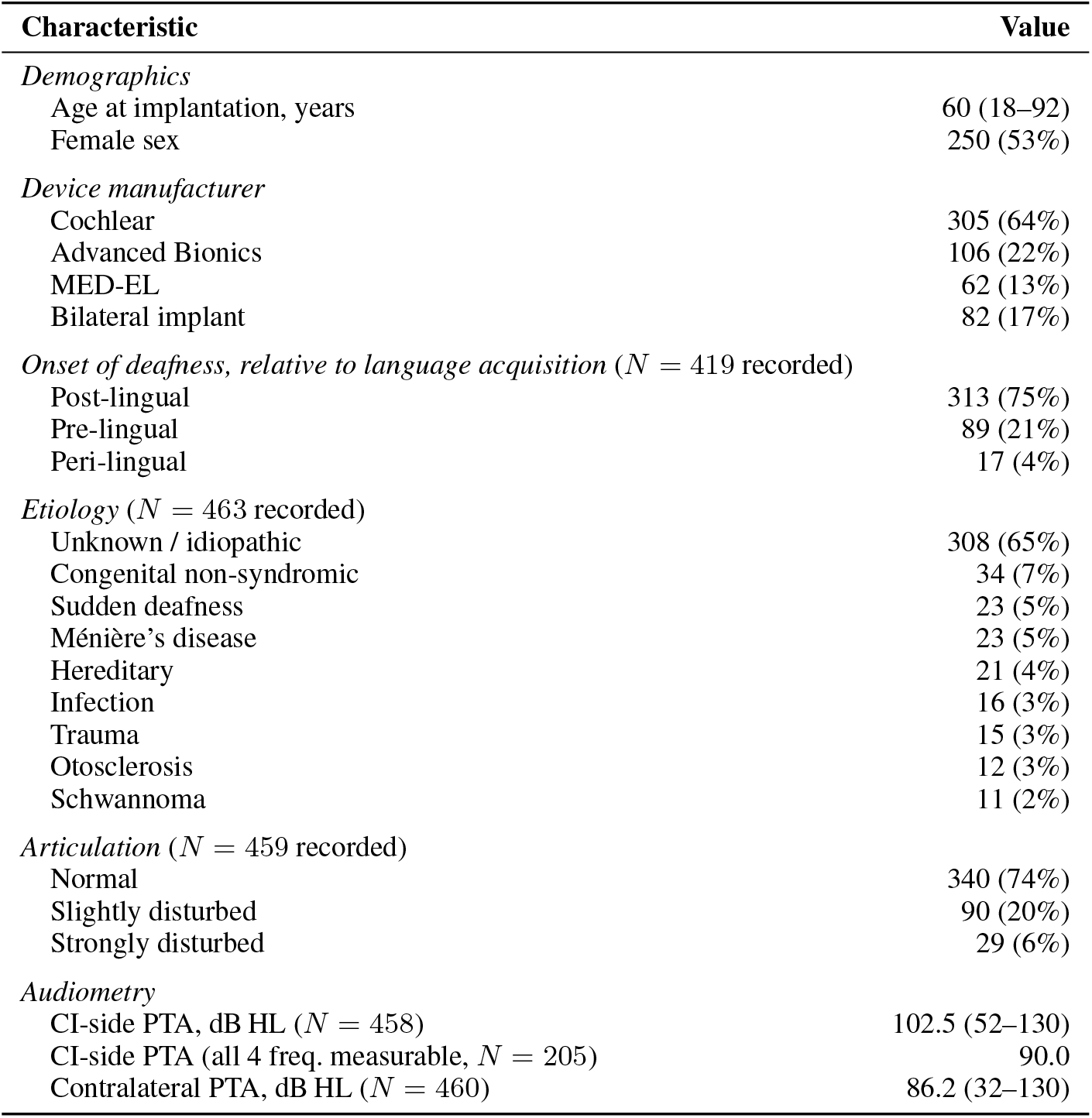
Demographic and clinical characteristics of the study cohort (*N* = 473 adults, SSD excluded). Continuous variables are reported as median (range); categorical variables as *N* (%), with percentages computed on the patients for whom the variable was recorded. Not all variables are complete: onset of deafness was recorded for 419 patients, articulation for 459, and etiology for 463 (the nine etiologies listed account for 463 patients; rarer causes are not shown individually). Articulation is transcribed from the pre-operative patient questionnaire and is therefore self-rated at that stage; it is updated by the audiologist post-operatively where necessary. PTA = four-frequency pure-tone average (500, 1000, 2000, 4000 Hz); unmeasurable thresholds are set to 130 dB HL. Onset categories refer to the timing of hearing loss relative to language acquisition, not to hearing status at implantation. All patients in this cohort were implanted as adults.

| Characteristic | Value |
| --- | --- |
| *Demographics* |  |
| Age at implantation, years | 60 (18–92) |
| Female sex | 250 (53%) |
| *Device manufacturer* |  |
| Cochlear | 305 (64%) |
| Advanced Bionics | 106 (22%) |
| MED-EL | 62 (13%) |
| Bilateral implant | 82 (17%) |
| *Onset of deafness, relative to language acquisition ($N = 419$ recorded)* |  |
| Post-lingual | 313 (75%) |
| Pre-lingual | 89 (21%) |
| Peri-lingual | 17 (4%) |
| *Etiology ($N = 463$ recorded)* |  |
| Unknown / idiopathic | 308 (65%) |
| Congenital non-syndromic | 34 (7%) |
| Sudden deafness | 23 (5%) |
| Ménière’s disease | 23 (5%) |
| Hereditary | 21 (4%) |
| Infection | 16 (3%) |
| Trauma | 15 (3%) |
| Otosclerosis | 12 (3%) |
| Schwannoma | 11 (2%) |
| *Articulation ($N = 459$ recorded)* |  |
| Normal | 340 (74%) |
| Slightly disturbed | 90 (20%) |
| Strongly disturbed | 29 (6%) |
| *Audiometry* |  |
| CI-side PTA, dB HL ($N = 458$) | 102.5 (52–130) |
| CI-side PTA (all 4 freq, measurable, $N = 205$) | 90.0 |
| Contralateral PTA, dB HL ($N = 460$) | 86.2 (32–130) |

#### 2.1.1 Speech Tests

The speech test battery comprised five tests, all administered in quiet at 65 dB SPL in the free field, each ear tested separately and aided: pre-operatively with the patient’s own hearing device where applicable, post-operatively with the cochlear implant alone. All tests were administered using the MACarena computerised test platform (Lai and Dillier, 2002). Three tests (monosyllable word recognition [WRS], vowel identification, consonant identification) are mandatory across all five Swiss CI centres and were administered pre-operatively and at post-operative follow-up visits; two additional tests (voice discrimination, number recognition) are administered routinely at University Hospital Zurich. WRS (Freiburger monosyllables), the primary outcome measure, is an open-set test in which the patient hears and repeats a list of 20 monosyllabic German nouns; it is scored as the percentage of words correctly identified and is a mandatory outcome measure across all five Swiss CI centres (Hahlbrock, 1953; Brand et al., 2014). A second equivalent WRS list was presented in the same session and is used here to estimate within-session test reliability (Supplementary Material, Table S8). The four remaining tests are described in Supplementary Material, Section S8. Not all patients had completed all five speech tests or had reached all follow-up intervals at the time of data extraction. Analyses therefore used different, partially overlapping subsets of the cohort. The four pre-operative speech tests other than WRS are used as predictors. For the trajectory analyses, post-operative WRS scores were grouped into four intervals (0–6, 6–12, 12–24, and *>*24 months) and used as outcome measure or predictors depending on the prediction task.

#### 2.1.2 Pre-Operative Features

The CICHDB extract contained 257 recorded items per patient. Items with more than 50% missingness were excluded, yielding a pool of 57 pre-operative features used as predictors for the analyses. Pre-operative features included audiometric thresholds for both ears, aided free-field thresholds, the four pre-operative speech tests other than WRS, demographic, clinical, socioeconomic and communication variables (full list in Supplementary Material, Section S8). Feature sets of varying sizes were constructed from this pool of 57 pre-operative features to test sensitivity to feature selection (Section 2.2).

### 2.2 Individual Prediction of CI Speech Outcome

We tested whether pre-operative data can predict individual post-operative monosyllable word scores (WRS) and performed a systematic comparison spanning prediction algorithms, feature sets, imputation strategies and validation procedures.

Seven prediction algorithms (Linear regression; Ridge regression (Hoerl and Kennard, 1970); Random Forest (Breiman, 2001); Gradient Boosting (Friedman, 2001); XGBoost (Chen and Guestrin, 2016); LightGBM (Ke et al., 2017); and the foundation model TabPFN (Hollmann et al., 2025)) were applied to feature sets of pre-operative variables to predict WRS scores. As TabPFN is a pre-trained foundation model for tabular data, it was fine-tuned on the dataset following the same cross-validation folds as for the other algorithms and described below (Supplementary Material, Section S3).

Feature sets used a subset of 3, 7, 17 or all 57 pre-operative features with pairwise interaction terms, and were constructed using data-driven selection via adaptive best-subset selection (ABESS) and Lasso regularisation. Pre-operative WRS scores were not included among the predictors in the primary analyses, because paired pre- and post-operative WRS scores were only available for a smaller subsample of *N* = 264 and including pre-operative WRS score as an additional predictor on the paired sample did not improve prediction performance with the best model TabPFN (see “Sensitivity analyses” and Supplementary Material, Section S4). Missingness in pre-operative predictors ranged from 25% to 40%; post-operative speech outcomes were not imputed. Median imputation was used for the primary analyses (Supplementary Material, Section S8).

Results are reported under *k*-fold cross-validation (*k* = 10 for the primary *N* = 378 sample, *k* = 5 for the paired *N* = 264 sensitivity analysis), and compared to in-sample results which are prone to over-fitting but have been used in previous studies (van de Velde et al., 2021; Steyerberg, 2019). Robustness to cross-validation methodology was probed with a 70/30 hold-out split, 5-fold and leave-one-out cross-validation, and a temporal split training on patients implanted before 2022 (Supplementary Material, Section S8) and testing on those implanted afterwards. We computed 90% prediction intervals by split conformal prediction (Vovk et al., 2005; Lei et al., 2018): the model is fitted on a training split, absolute residuals are computed on a held-out calibration split, and the interval half-width is the 90th percentile of those residuals. The intervals are therefore distribution-free and have marginal 90% coverage by construction.

Prediction performance was assessed with cross-validated *R*^2^ (proportion of variance explained; can be negative when the model underperforms the sample mean) and mean absolute error (MAE; the average absolute difference between predicted and true WRS score, in percentage points).

#### 2.2.1 Analysis of Predictor-Outcome Relationship

If pre-operative features cannot predict post-operative scores reliably, there are two possible explanations: there is not sufficient pre-operative information about speech outcome per se, or the relationship of pre-operative information with speech outcome changed after surgery. The two can be separated, because every patient is measured twice on the same speech tests, once before surgery and once after. A prediction model uses all the features collectively to achieve its best possible prediction but does not specify why it fails. Here we analysed pre-operative features one at a time, to assess predictive relationships with speech outcome before and after CI surgery and to establish whether a change in predictive information is general or affects only some pre-operative features.

We first analysed this for pre-operative features measured on a continuous scale, such as PTA, where a correlation indicates whether scores rise or fall as the variable increases or decreases. We found that pre-operative features correlated more with WRS before than after CI surgery. As an example, pre-operative CI-side PTA correlated with pre-operative WRS score (Spearman *ρ* = −0.31, *p* < .001, N = 262) and with post-operative WRS score (*ρ* = −0.15, *p* = .005, N = 366; bootstrap difference *p* = .015), but at a significantly lower level (*p* = .038). The other three speech tests showed the same pattern, with pre-operative correlations falling to |*ρ*| < 0.20 post-operatively. This preliminary analysis indicated that the pre-operative information for speech outcome prediction is changed (disrupted) after CI surgery. A correlation, however, only tracks whether scores go up or down, and only works for variables measured on a continuous scale. It cannot be used for categorical pre-operative features such as articulation, onset of deafness or language history.

Instead of using correlation, we therefore compared score distributions between feature-specific groups. For each pre-operative feature, patients were split into groups according to their characteristic value: articulation, for example, was split into three groups (normal, slightly disturbed, strongly disturbed), and PTA was split into several groups with different ranges of hearing loss. We then assessed the score distributions of groups to determine the value of a given feature for predicting speech outcome: if patients with mild and profound hearing loss have similar speech scores, knowing which group a patient belongs to does not help in outcome prediction. In this manner, we analysed score distributions and their distances before and after CI surgery features. The distances between distributions were measured with the Wasserstein distance (*W*_1_; Peyré and Cuturi 2019), which compares the whole distributions rather than only their averages, is expressed in the same units as the score, and works whether a variable defines two groups or several. The change in *W*_1_ from before to after CI surgery was statistically compared by permutation testing (5,000 label permutations; Supplementary Material, Section S2). A lower value for *W*_1_ is obtained when distributions overlap whereas a higher value is obtained when distributions are more different.

### 2.3 Risk Classification for Poor CI Speech Outcome

We reformulated the prediction problem as a binary classification task, following recent work that has shifted from continuous outcome prediction toward identification of poor performers (Shew et al., 2025; Fraysse and James, 2019). Poor absolute outcome was defined as a post-operative WRS score below 40%, approximately the 20th percentile of the post-operative distribution in our cohort. There is no established threshold for poor cochlear implant outcome and the poor-performer category, while clinically recognised, is not sharply bounded (Moberly, 2016). We therefore adopt the 40% cutoff pragmatically, as a cohort-based operational definition rather than a diagnostic standard, verified by a threshold-sensitivity analysis (Supplementary Material, Table S5).

Classification was performed with logistic regression, Random Forest, and TabPFN on the full set of 57 pre-operative features, under 10-fold stratified cross-validation, and discrimination was reported as the area under the ROC curve (AUC): the probability that a randomly chosen poor performer is assigned a higher predicted risk than a randomly chosen non-poor performer, where 0.5 is chance and 1.0 is perfect separation. Because AUC is invariant to class prevalence, the three models remain comparable despite differing in how they handle class imbalance (logistic regression and Random Forest used balanced class weights; TabPFN was applied to the data as provided). Logistic regression is reported alongside the non-linear models because it yields interpretable odds ratios for individual risk factors. Model settings are given in Supplementary Material, Section S3.

To identify the features with the most information for risk classification, univariate odds ratios with 95% confidence intervals were computed for each pre-operative variable. Bonferroni correction was applied over the 49 features for which a univariate model of poor outcome could be fitted. This is fewer than the 57 features in the pool, because nominal categorical variables with several unordered levels and variables with a near-constant distribution do not yield a single odds ratio. The corrected threshold is therefore 0.05*/*49 = 0.00102. For audiometric and aided-field variables, unmeasurable thresholds were excluded rather than imputed (Supplementary Material, Table S3). Permutation importance identified the most informative features in the Random Forest and TabPFN models, and the classifiers were retrained on audiometric, developmental-communicative, and other variable subsets separately to compare their AUC.

### 2.4 Trajectory Analysis for CI Speech Outcome

We examined whether early post-operative speech outcomes carry prognostic information for later speech outcomes. The six-month post-operative WRS score was added to the 57-feature pre-operative set as a single additional predictor, and TabPFN was applied to predict long-term post-operative WRS scores ( ≥ 12 months) under 10-fold cross-validation in comparison to the prediction without post-operative WRS score included. The same comparison was repeated as a binary classification: the models were trained to identify poor long-term outcome (WRS < 40%) from the pre-operative features alone and with the six-month post-operative score added, and discrimination was again reported as AUC.

To characterise post-operative trajectory patterns, a clustering analysis was performed on patients with at least three serial post-operative WRS scores spanning early (0–6 months) and late (12–24 months) intervals (*N* = 136). *k*-means clustering was applied to the standardised score vectors with an optimal number of clusters selected by silhouette analysis. Trajectory clusters were then used as classification targets to determine whether post-operative trajectories in speech performance (rather than absolute scores or poor performances) can be predicted from pre-operative data. Two prediction models (Random Forest and TabPFN) were applied to predict cluster membership from the 57 pre-operative features under 10-fold stratified cross-validation. The prediction of cluster membership, unlike the clustering itself, was treated as exploratory given the unbalanced four-class setting.

## 3 Results

### 3.1 Outcome Variability

Post-operative WRS scores (*N* = 378) spanned the full range from 0 to 100%, with a group mean of 62.3% (±26.0). Mean scores differed by onset of deafness (post-lingual 66%, N = 257; peri-lingual 63%, N = 12; pre-lingual 52%, N = 63), by articulation quality (normal 66%, N = 284; slightly disturbed 49%, N = 64; strongly disturbed 45%, N = 20), and across CI-side PTA quartiles (mildest, 55–90 dB HL: 70%, N = 95; profound, 114–130 dB HL: 57%, N = 90; Spearman *ρ* = −0.15, *p* = .005). By etiology, mean WRS ranged from 74% in Ménière’s disease (N = 12) to 33% in infection (N = 5). Age at implantation showed no association with outcome (*r* = 0.02, *p* = .69). Eight adults scored 0% on WRS; the same patients reached up to 100% on number recognition, up to 100% on voice discrimination and up to 58% on vowel identification, and three had documented anatomical abnormalities (gusher at insertion, obliteration of the round-window niche and basal scala tympani, and sclerotic petrous bone).

### 3.2 Prediction of CI Speech Outcomes

No prediction algorithm predicted individual post-operative WRS at a useful level when tested on held-out data (Table 2). Cross-validated *R*^2^ was at or below zero for most algorithms and feature-set combinations, while in-sample *R*^2^ reached 0.86 for Random Forest and 0.94 for XGBoost due to likely overfitting of the data. Linear models sat near zero on the small feature sets (+0.01 at 3 features) and fell to −0.27 at 57 features. The highest cross-validated value in the grid was *R*^2^ = 0.22, reached by TabPFN on the 57-feature set. Mean absolute error ranged from 19 to 24 percentage points across models, so even the best model’s prediction was on average about 19 points away from the patient’s actual score. Ninety per cent conformal prediction intervals spanned ±41 percentage points around each predicted score (± 37 for TabPFN at 57 features): a patient predicted at 60% has an interval from 19% to 100%. Empirical coverage matched the nominal target.

**Table 2:** Prediction performance for individual post-operative WRS (*N* = 378 adults; with the four pre-operative speech tests other than WRS included as predictors, see Section 2.1.2). For each algorithm and feature-set size, in-sample *R*^2^ (training-pass fit, including overfitting) is reported alongside the cross-validated (CV) *R*^2^ (10-fold cross-validation, pooled out-of-fold predictions, averaged across 3 random seeds). Bold marks the best cross-validated value in each column; in-sample values are reported only to show the potential for overfitting and should not be read as performance.

| Algorithm | 3 features |  | 7 features |  | 17 features |  | 57 features |  | MAE<br>(57 feat.) |
| --- | --- | --- | --- | --- | --- | --- | --- | --- | --- |
|  | In-s. | CV | In-s. | CV | In-s. | CV | In-s. | CV |  |
| Linear regression | 0.03 | +0.01 | 0.03 | +0.00 | 0.06 | -0.03 | 0.21 | -0.27 | 24 |
| Ridge regression | 0.03 | +0.01 | 0.03 | +0.00 | 0.06 | -0.03 | 0.20 | -0.24 | 24 |
| Random Forest | 0.76 | -0.23 | 0.84 | -0.11 | 0.86 | -0.05 | 0.86 | -0.03 | 22 |
| Gradient Boosting | 0.39 | -0.10 | 0.47 | -0.05 | 0.59 | -0.06 | 0.72 | -0.04 | 22 |
| XGBoost | 0.89 | -0.58 | 0.91 | -0.40 | 0.93 | -0.21 | 0.94 | -0.19 | 24 |
| LightGBM | 0.38 | -0.17 | 0.55 | -0.18 | 0.75 | -0.21 | 0.94 | -0.09 | 23 |
| **TabPFN (fine-tuned)** | 0.57 | -0.01 | 0.61 | 0.16 | 0.66 | 0.16 | 0.69 | 0.22 | 19 |

The same pattern of results was robustly observed for various validation approaches and secondary analyses. A 70/30-training/testing split, 5-fold and leave-one-out cross-validation, and a temporal training/testing split (training before 2022, testing from 2022 onward) all returned *R*^2^ at or below zero, the temporal split being the lowest. Results were unchanged under Lasso feature selection, alternative outcome formulations (log, arcsine-square-root, change score), and inclusion of pre-operative WRS in the paired sample (*R*^2^ ≈ 0.10). On random subsamples of 25%, 50%, 75% and 100% of the cohort, tree-ensemble *R*^2^ rose from -0.08 at N = 94 to +0.04 at N = 189 and then stayed flat (+0.03 at N = 283 and N = 378). TabPFN improved with sample size and reached *R*^2^ = 0.22 at the full cohort. Using a smaller subsample of 284 participants with WRS measured at least 12 months (rather than 6 months) after CI activation did not improve *R*^2^. Further details are reported in Supplementary Material, Section S4. The same pre-operative features predicted the *pre*-operative WRS score much better with TabPFN reaching a cross-validated *R*^2^ of 0.52, and of 0.48 when the other pre-operative speech tests were excluded, against 0.22 for the post-operative score.

### 3.3 Changes to Predictor-Outcome Relationship

The predictor-outcome relationship changed after CI implantation for several pre-operative features. For the audiometric, aided-field and pre-operative speech features, the feature-specific groups scored in the same range after implantation. PTA as example had patients in the mildest and most profound hearing-loss groups scoring differently before implantation but their score distributions overlapped strongly after implantation. Across all these features, the average *W*_1_ distance between their groups fell by about seven tenths, from 21.5 to 6.0 percentage points in speech scores. This was the case while patients within each feature-specific group still differed in terms of speech outcomes as much as before (mean SD of 27.4 compared to SD of 25.0). There was a much larger overlap between feature-specific groups, but not much less variability among patients.

For other features the predictor-outcome relationship changed after CI implantation, but in the opposite direction. Onset of deafness had a *W*_1_ distance of 3.6 before and a much larger *W*_1_ distance of 17.1 percentage points after implantation (*p* < .001), while articulation slightly increased from 18.3 to 19.7 (*p* = .77), which was the largest post-operative distance of any feature in the pool. Again there was a comparable variability among patients within the feature-specific groups, but there was still a large distance between distributions after implantation.

The features with converging and overlapping distributions after implantation were the audiometric and speech measures, while developmental and communicative features retained or increased their distances between feature-specific groups. There were no outliers as none of the distributions of the 33 audiometric, aided-field or pre-operative speech features moved apart after implantation. All seven features that had less overlapping distributions were developmental or communicative. The pre-operative speech tests, the measures most similar to the outcome itself, lost the most predictive power: their mean distance fell from 32.3 to 5.2 percentage points, against 13.2 to 6.1 for the audiometric and aided-field thresholds. The features with less overlap in distributions started from little separation (mean 6.1 before implantation) and ended up separating patients more than any other pre-operative measure (mean 15.0). Distances before and after implantation, with permutation *p*-values, are given for every feature in Supplementary Material, Table S9 and Figure S3. Of the 46 features for which a Wasserstein distance could be computed, 23 converged, 7 moved apart and 16 showed no significant change from pre- to post-implantation. Figure 1 shows both patterns of convergence and separation, for onset of deafness and PTA of the implanted ear. Further details on this analysis are reported in Supplementary Material, Section S2.

**Figure 1:**
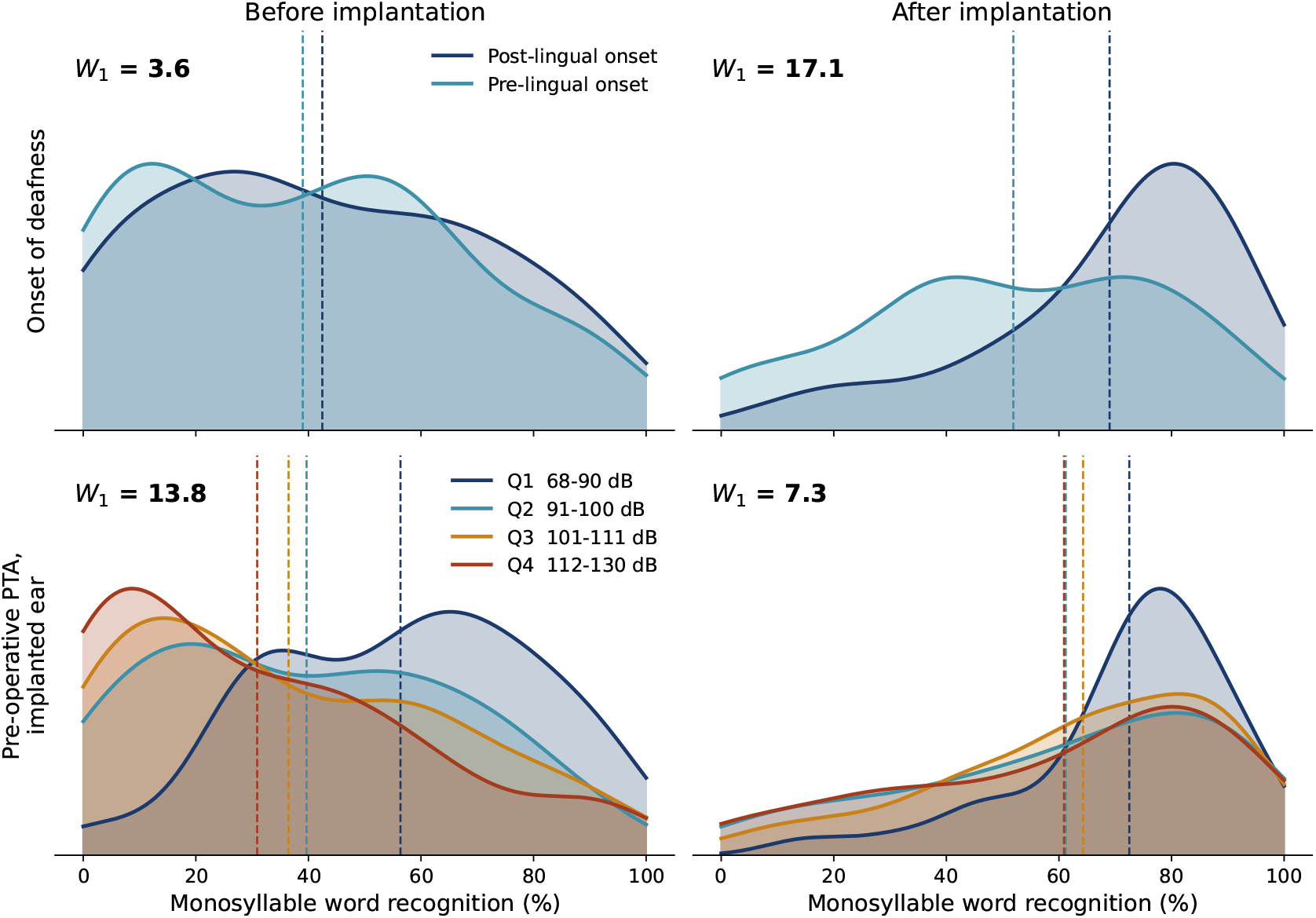
Distribution of monosyllable WRS by group, before and after implantation, for two pre-operative features: onset of deafness (top) and pre-operative PTA of the implanted ear in quartiles (bottom). Each curve is the score distribution of one feature-specific group; dashed lines mark group means. *W*_1_ is the mean Wasserstein distance between the groups’ distributions in that panel, in percentage WRS: it is large when the groups occupy different parts of the WRS score range and small when they overlap.

### 3.4 Identifying Patients at Risk of Poor Outcomes

Poor absolute outcome was defined as a post-operative WRS score below 40%, approximately the 20th percentile of the post-operative distribution. Of the 378 adults, 75 (19.8%) met this criterion, distributed across the full pre-operative score range. TabPFN, fine-tuned for binary classification, discriminated these patients with AUC = 0.71. Logistic Regression with balanced class weights achieved a slightly lower AUC = 0.67. Discrimination performance was stable across poor-performance thresholds from 30% to 50%. The model separated high-risk from low-risk patients, but the assigned probabilities were too high: in the highest-risk quartile it predicted a 79% chance of poor outcome whereas 33% was observed in the data (Supplementary Material, Table S6).

Articulation quality was the only variable to survive Bonferroni correction in univariate logistic regression (OR = 2.55, 95% CI [1.70, 3.81], *p* < .001; corrected threshold 0.05*/*49 = 0.00102). The next strongest nominal associations were learned profession and education, followed by sign language history, multilingual home, onset of deafness and family history of CI use, but none of these reached the corrected threshold. Several audiometric, aided-field and pre-operative speech measures reached nominal significance (*p* < .05) without surviving correction, and age showed no univariate association (OR near 1.00). Permutation importance from TabPFN also ranked articulation first, approximately 2.5× above the next variable, followed by age, mid-to-high-frequency thresholds and further developmental and communicative variables. Further details are provided in Supplementary Material, Table S9, with tests for each feature.

When restricted to the 35 audiometric, aided-field and pre-operative speech variables, Logistic Regression reached AUC = 0.529. The 6 developmental and communicative variables reached AUC = 0.641 and the 5 demographic and clinical variables reached AUC = 0.650. Random Forest and TabPFN reproduced the same ordering (TabPFN: 0.585, 0.654 and 0.666; Figure 2, Supplementary Material, Table S10). Cross-validated *R*^2^ for the same subsets remained below 0.10 in every case and did not reproduce this ordering.

**Figure 2:**
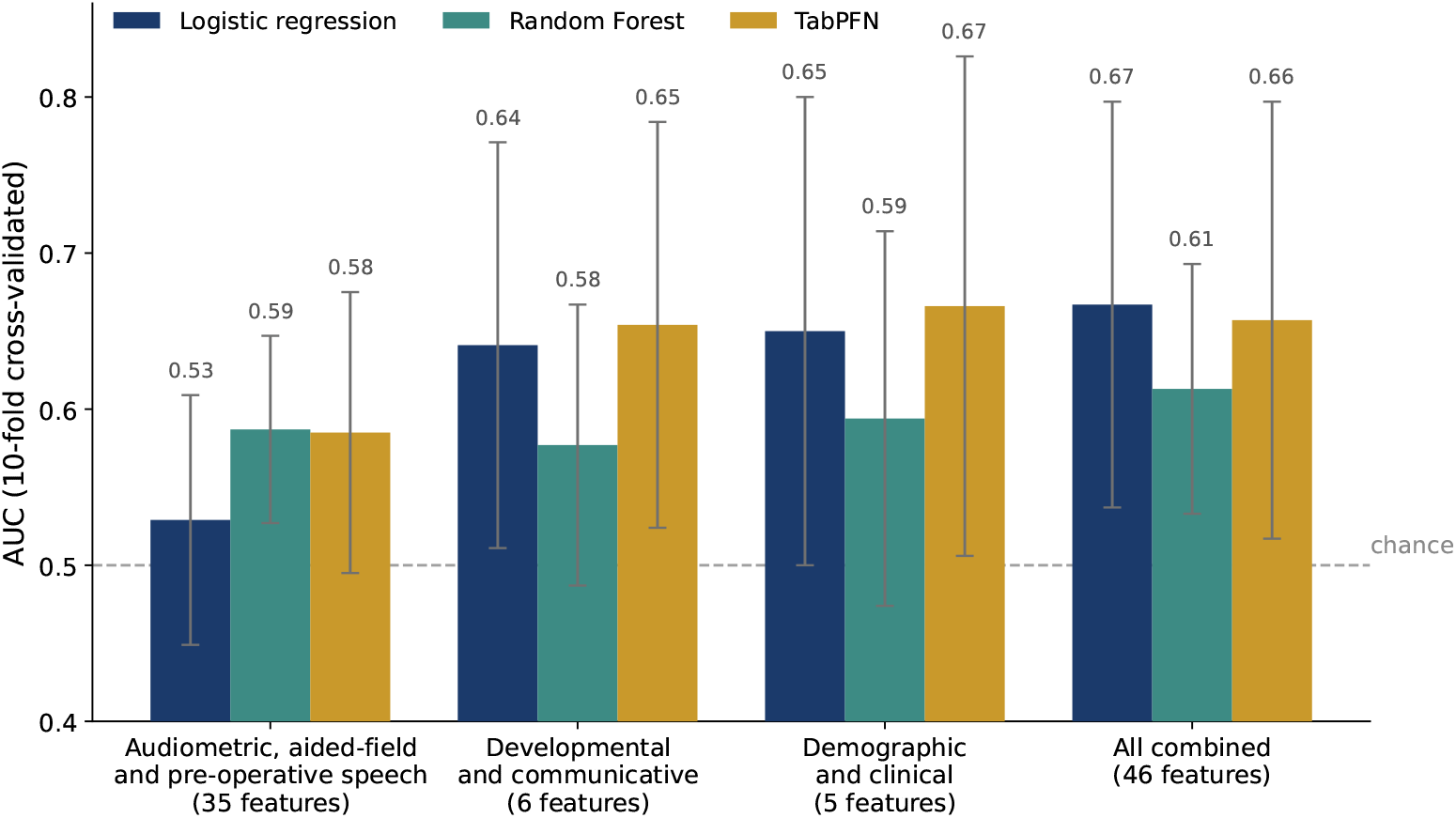
Cross-validated AUC for poor outcome risk prediction (WRS < 40%, *N* = 378) by feature subset and algorithm (Logistic Regression, Random Forest, TabPFN). Pre-operative feature subsets used from left to right: 35 audiometric, aided-field and pre-operative speech variables, as often used in the CI prediction literature; 6 developmental and communicative variables; 5 demographic and clinical variables; and all 46 variables combined. Error bars indicate one standard deviation across 10 cross-validation folds. Dashed line: chance level (AUC = 0.5).

**Figure 3:**
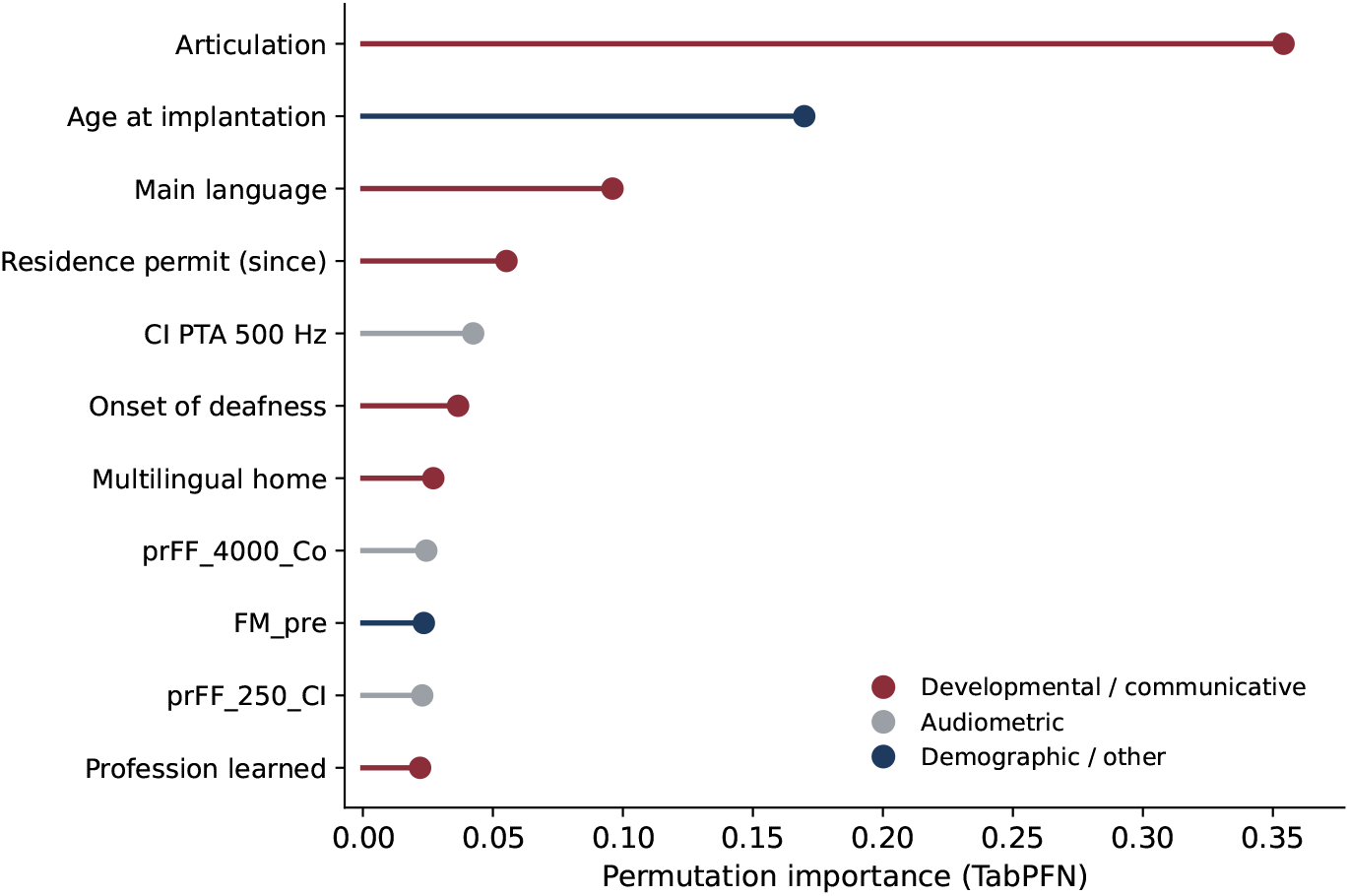
TabPFN permutation importance for the 12 best-performing pre-operative predictors of poor outcome (WRS < 40%, *N* = 378)

### 3.5 Post-Operative Trajectories of Speech Outcomes

In the *N* = 188 adults with both a 0–6 month and a 12–24 month WRS score, cross-validated *R*^2^ for long-term WRS from the 57 pre-operative features alone was 0.03 (TabPFN). Adding the six-month post-operative WRS score as a 58th predictor raised *R*^2^ to 0.48. The six-month score alone, without any pre-operative variable, reached *R*^2^ = 0.36. Pre-operative WRS correlated with long-term WRS at *r* = 0.15 and with the six-month score at *r* = 0.69. For comparison, the within-session correlation of WRS was *r* = 0.92 between the two word lists (*N* = 378) and can be seen as an upper bound for explained variance (*R*^2^ = 0.85 for ICC = 0.92). When considering this upper bound, the six-month WRS score therefore reached 42% of the total explainable variance, indicating that the remaining variance reflects a genuine change in speech performance after six months rather than measurement noise. Pre-operative features could also not explain the residual variance left by the six-month score (cross-validated *R*^2^ ≤ 0 for all algorithms). Relaxing the inclusion criterion to any measurement at 12 months or later raised the sample to *N* = 207 and left the result unchanged (*R*^2^ = 0.49 from the six-month score, *R*^2^ = 0.00 from pre-operative WRS). The same was found for risk classification. Using the six-month post-operative score in addition to the pre-operative features, poor long-term outcome was identified with AUC = 0.84, against AUC = 0.71 from pre-operative data alone.

The 188 patients with serial measurements scored higher than the remainder of the cohort (mean WRS 68.8% versus 55.9%; poor outcomes 8.6% versus 30.9%; both *p* < .001) and had lower outcome variance (SD 21.5 versus 28.5 percentage points). However, they did not differ on several other pre-operative variables tested (age, CI-side and contralateral PTA, pre-operative WRS, articulation, onset of deafness; all *p >* .16).

*k*-means clustering of standardised serial WRS scores in the *N* = 136 adults with at least three measurements gave *k* = 4 by silhouette analysis (Figure 4): Early Responders, who plateau quickly (50%, *N* = 68); Late Responders, who continue to improve past six months (19%, *N* = 26); Non-Responders, whose scores change little (21%, *N* = 29); and Decliners, whose post-operative scores fall below their pre-operative WRS score (10%, *N* = 13). The clusters differed sharply in pre-operative score, from a mean of 29% in Early Responders to 80% in Decliners, but with a smaller difference for long-term WRS, with 76% and 50%, respectively. Random Forest and TabPFN classifiers were trained to predict cluster membership from pre-operative features and reached a mean accuracy of 51%, against a majority-class baseline of 50% and a proportional-chance baseline of 34%. Adding the six-month post-operative score to the pre-operative features raised four-class accuracy from 57% to 71%. However, this gain is partly definitional, since the trajectory types were defined by the change from pre-operative WRS to post implantation WRS. For example for discriminating Early Responders from the rest using pre-operative and six-month WRS alone reached AUC 0.97 with TabPFN.

**Figure 4:**
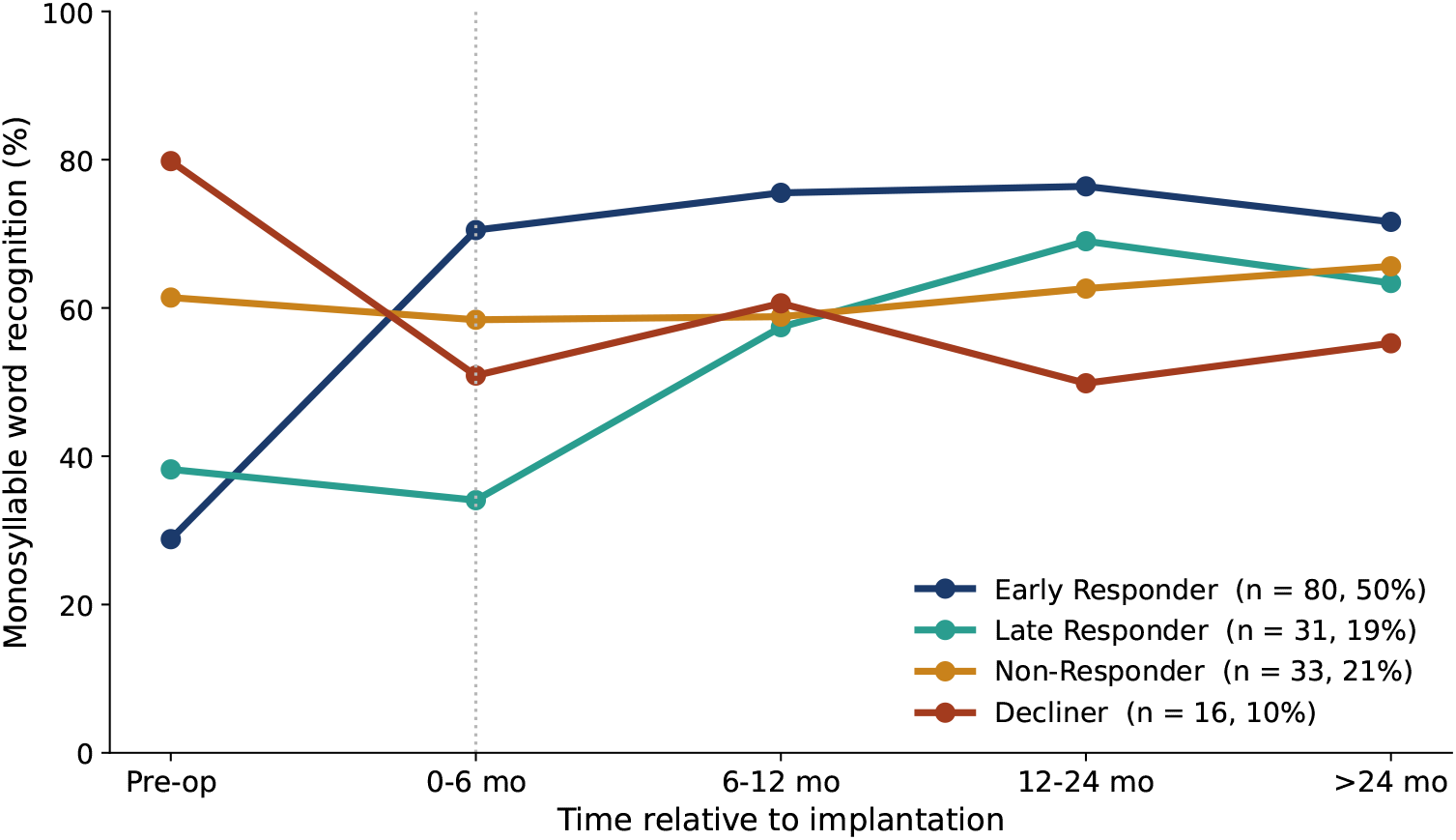
Average WRS across follow-up intervals for the four trajectory clusters (*N* = 136; *k*-means on standardised serial scores, *k* = 4 by silhouette analysis). Early Responders plateau quickly, Late Responders continue improving past six months, Non-Responders change little, and Decliners end below their pre-operative score. The dotted line marks the six-month WRS score after implantation.

## 4 Discussion

We confirm that pre-operative variables collected in clinical practice fail to predict CI speech outcomes across seven prediction algorithms, four feature-sets, five imputation strategies, and six validation procedures. Cross-validated *R*^2^ for individual post-operative WRS reached a maximum of 0.22 with the most capable prediction model, a recent foundation model pre-trained on vast datasets and fine-tuned on the training data in this study (Hollmann et al. 2025). This is consistent with the modest predictive performance of the largest multicentre studies, whether expressed as variance explained (*R*^2^ around 10% by Blamey et al. 2013 and 8 to 11% by Anthonisen et al. 2026) or as prediction error (mean absolute error of 17 to 22 percentage points by Shafieibavani et al. 2021 and Demyanchuk et al. 2025), although direct comparisons are complicated due to the different cohorts, outcome measures and feature sets used. Because this limited prediction performance occurs across various datasets (including a subsample of 284 participants with WRS measured at least 12 months after CI activation) and prediction approaches (various algorithms compared here and in previous studies), it likely reflects the limited information contained in the pre-operative data with respect to post-operative CI speech outcomes, and not a limitation of the prediction models and algorithms used. This interpretation is in line with recent observations that pre-operative measures do not capture auditory-nerve function directly (Bartholomew et al., 2024), as they rely on acoustic hearing function, and so do not appropriately reflect factors important for electric hearing function. The obvious difference in hearing function before and after CI implantation may explain why the pre-operative data related to acoustic hearing function only has limited information for CI outcome prediction. Further evidence for this assumption comes from the results when applying the same models to predict the pre-operative speech score instead of the post-operative one. Predicting pre-operative WRS from the same pre-operative features led to a cross-validated *R*^2^ of 0.52 with the best model TabPFN, and only dropped slightly to 0.48 when using only audiometry features without any pre-operative speech test data. This shows that the pre-operative features contain highly useful information about speech recognition outcomes in the acoustic hearing domain, but seem to lose that information once hearing is mediated by the CI.

It is important to note that here was a large difference in prediction performance between in-sample and cross-validated testing data for all the algorithms. Several previous studies only reported in-sample prediction results which could be misleading. For example, ensemble algorithms (Random Forest, XGBoost, LightGBM) predicted (or “learned”) the in-sample data almost perfectly (in-sample *R*^2^ up to 0.94) but failed to predict unseen data under cross-validation or similar approaches that hold-out data for the testing. Overfitting is very likely to occur when prediction algorithms are trained and tested on the same data, leading to hugely inflated performance at the cost of generalisation to unseen, realistic test data.

In an attempt to explain the limited prediction performance, we used a distributional analysis of the 57 pre-operative variables (Supplementary Material, Section S2) and showed that distributional differences before CI implantation largely collapse afterwards in respect to CI speech outcomes: the implanted ear’s residual hearing, the contralateral ear’s residual hearing, and the aided thresholds stratify patients well before surgery but poorly afterwards, likely because CI implantation bypasses the peripheral factors that these measures index. The variables that instead retain or increase their distributional differences in respect to CI speech outcomes are developmental and experiential variables including onset of deafness, language history, and learned profession. These variables index higher-level properties of auditory and language processing that do not change with CI implantation. Some of these variables, such as main language and residence-permit status, are proxies for socio-linguistic and educational background rather than clinical characteristics and the modest information they carry may therefore reflect social and linguistic factors rather than auditory or cognitive ones. Articulation quality and several other variables showed no significant change in distributional difference before and after CI implantation. As discussed before, variables that describe the acoustic hearing system seem unable to determine performance once electrical hearing is used, but different kinds of data (neural, cognitive, and biological) may overcome this limitation of routine clinical pre-operative variables. A recent prospective study illustrates such data: spectro-temporal ripple discrimination measured shortly after activation predicted speech-in-noise recognition one year later (*r* = 0.51) and outperformed the Freiburger word test (Erb et al., 2026). One caveat applies here: pre-operative WRS scores are obtained in best-aided condition with both ears while post-operative WRS scores are obtained with the CI ear alone, so there are more differences in listening condition as well as in transduction type.

The same phenomenon explains both the limit on continuous prediction and the partial success of binary risk classification: the variables that conventional models often rely on are also the variables that CI implantation compresses, while the variables that survive support coarse risk classification more readily than continuous prediction. Because binary risk rests on differences in subgroup means rather than on individual-level precision, it is captured comparably by simple and flexible models: the advantage of the foundation model in continuous-score regression largely disappears for the coarser classification target (Figure 2).

Reformulated as binary classification of poor outcome, pre-operative data become partially informative (AUC ≈ 0.71), driven by articulation and other developmental and communicative variables: sufficient for risk-informed counselling, but far from individual prediction, and no better than chance when restricted to audiometric variables. The clinically actionable information on individual outcome instead arrives after surgery, where a single six-month score raises long-term *R*^2^ from 0.03 to 0.48. This mirrors prospective evidence from Peng et al. (2026) that post-operative psychophysical and cortical measures explain outcome variance that pre-operative history does not, and supports reframing CI outcome prediction as a longitudinal rather than cross-sectional problem. It should be noted that the cross-session correlation between six-month score and long-term score is still considerably smaller than the within-session correlation of the same test (*r* = 0.69 versus *r* = 0.92). This indicates that the test is reliable and that some variance of the long-term outcome is determined after the first six months as expected.

The four trajectory types identified in this longitudinal subsample (*N* = 136, large for serial CI outcome data) refine this picture: early and late responders, non-responders, and a small group who decline below baseline. Trajectory type cannot be anticipated from pre-operative features but is evident from the early post-operative course itself, so the question whether a patient will achieve good CI speech scores is currently unanswerable before surgery yet emerges by six months post surgery in the early phase of acclimatization.

The findings translate into two clinical stages, before and after CI surgery, by guiding pre-CI counselling and post-CI rehabilitation. Patient counselling should take into account that individual post-operative scores can currently not be predicted from pre-operative data, because a 90% prediction interval spans nearly the full outcome range.

Disturbed articulation, together with developmental and communicative variables (onset of deafness, multilingual home environment, family history of CI use), partially identify patients at risk of poor absolute outcome (AUC ≈ 0.71). These factors can somewhat inform expectation management and rehabilitation planning, but not surgical eligibility decisions. By six months post-implantation, the post-operative score becomes substantially more prognostic of long-term outcome than any pre-operative variable (*R*^2^ = 0.48 with one post-operative score added to the 57-feature set, versus 0.03 from pre-operative features alone). Risk identification improves in the same way: poor long-term outcome is identified with AUC = 0.84 from the six-month battery, against 0.71 from pre-operative data alone. The six-month visit is the inflection point at which intensified rehabilitation, remapping, or additional assistive devices should be considered for optimizing rehabilitation for patients who are not yet receiving the expected CI benefit.

Several limitations should be noted. The current analysis is single-centre and retrospective. While the 57 pre-operative features are standardised nationally and broadly representative of routine European practice, the AUC and *R*^2^ values obtained here may not transfer to other CI centres with different test batteries, candidacy criteria or follow-up schedules. The trajectory clusters and the six-month prognostic window are descriptive of the present cohort and require prospective replication before they are deployed as clinical decision aids. For risk predictions, we set a 40% threshold to define poor outcome corresponding approximately to the 20th percentile in this cohort, but this is a pragmatic cutoff and arbitrary threshold which may shift under alternative definitions of poor outcome or with other cohorts. Articulation quality, the dominant pre-operative risk predictor for poor outcome, is a coarse categorical rating transcribed from the patient questionnaire before surgery and therefore self-rated at the pre-operative stage, and it is likely related to pre-lingual onset of deafness, language exposure and other cognitive-linguistic factors. It should be interpreted as a proxy for a patient’s communicative and developmental history, not as an independent cause or factor of poor outcome. The monosyllable WRS test’s psychometric properties constrain how well it can be predicted. WRS scores are measured in steps of 5 percentage points, and published test-retest differences for the Freiburger test range from ±13 to ±24 points. This implies a maximum achievable *R*^2^ of approximately 0.85, rather than 1.00, due to the within-session reliability in our cohort (ICC = 0.92; Supplementary Material, Table S8). When taking this into account, the best model still fails to explain more than a quarter of the explainable variance. Importantly, the main outcome measure is a monosyllable word-recognition score measured in quiet at somewhat variable intervals, consistent with national standards (Brand et al., 2014) and the CI prediction literature (Blamey et al., 2013; Lazard et al., 2012; Holden et al., 2013; Shafieibavani et al., 2021). Nonetheless it captures only a narrow slice of CI outcomes: speech understanding in noise, listening effort, sound quality, music enjoyment, quality of life and self-reported everyday function all matter to CI recipients and may only be loosely coupled to word recognition scores in quiet. This is supported by a meta-analysis on CI studies showing that speech-recognition scores correlate weakly, at best, with quality-of-life improvement (McRackan et al., 2018), and a previous study questioning whether word recognition scores measure what matters most to CI recipients (Moberly et al., 2018). Future development of implant-specific patient-reported measures may be warranted to address this challenge (McRackan et al., 2019). Furthermore, the non-availability of a post-operative WRS score excluded some patients from the dataset, which leads to a truncated dataset and potentially changed relationships between variables. An outcome measure that can be reliably obtained with all CI patients would be preferable to allow the inclusion of the whole population of CI patients. Therefore, the findings reported here only apply to the WRS outcomes and specific cohort.

### 4.1 Conclusion

Individual post-operative speech scores could not be reliably predicted from pre-operative data with seven prediction algorithms (best cross-validated *R*^2^ ≤ 0.22), including a powerful transformer-based foundation model. However, poor CI outcomes could be partially identified as a binary risk (AUC = 0.71; acceptable discrimination (Hosmer et al., 2013)) that was driven by developmental and communicative rather than audiometric variables. And adding the first post-operative speech score at 6 months to the predictor variables substantially raised prediction *R*^2^ to 0.48. The limited prediction performance seems to arise because cochlear implantation changes the predictive value of the variables routinely collected in clinics: speech score distributions separated by audiometric and pre-operative speech variables before surgery converged after surgery and lost their predictive value. However, prediction of the pre-operative score from the same variables was successful (*R*^2^ = 0.48 based on audiometric data only), indicating that the limitation was not related to insufficient pre-operative data quality. In contrast, the distributions separated by the remaining developmental and communicative variables diverged after surgery and could support coarser risk classification for poor outcomes rather than speech score prediction. These findings confirm that the limited information collected before CI implantation is unlikely to be sufficient for good prediction performance. CI outcome prediction may instead be seen as a longitudinal iterative process instead of a one-time prediction task. Our findings support a prediction-based two-stage framework to counsel for poor outcome risk before CI surgery and to track and forecast speech score trajectories after CI implantation to trigger intervention when needed via personalised rehabilitation, timely CI fittings, or additional assistive devices and services.

## Acknowledgements

MC was funded by the European Union’s Horizon Europe research and innovation programme under Marie Skłodowska-Curie Actions grant agreement No. 101275781 (REHEAR). TG was funded by the Swiss National Science Foundation (SNSF) [grant number 226368]. The authors thank Dr. Rahel Bertschinger, Dr. Marlies Geys and Dr.Michael Büchler.

## Data Availability Statement

The Swiss CI database is managed by University Hospital Zurich. Data sharing requests should be directed to the corresponding author and are subject to ethics approval and data protection regulations. Analysis code is available at https://github.com/mcampi111/CI_Outcome_Pred.

## Ethics Statement

This study was approved by the responsible cantonal research ethics committee and conducted in accordance with the Declaration of Helsinki. Approval number KEK-ZH-Nr. 2024-00881.

## Authors’ Contributions

M.C.: Conceptualization, data curation, formal analysis, investigation, methodology, software, visualization, original draft preparation, writing—review and editing.

A.H.: Resources, data curation, clinical interpretation, funding acquisition, writing—review and editing.

T.G.: Conceptualization, methodology, validation, funding acquisition, supervision, project administration, writing— original draft, writing—review and editing.

M.C. and A.H. confirm that they had full access to all the data in the study. All authors have read and agreed to the published version of the manuscript.

## List of Supplementary Material

Supplementary Material.pdf

