## Supplementary Information for "Prediction of Individual Speech Outcomes with Cochlear Implants: Limitations and Opportunities for Clinical Translation"

### SI Section S1: Overview of analyses

This section maps every analysis reported in the paper: the sample it uses, the predictors and the outcome, and where the result appears (Table S1).

Table S1: Every analysis reported in the paper, with the sample it uses, the predictors and outcome, the models applied, and where the result is reported. All subsets are drawn from the study cohort of $N = 473$ adults with bilateral hearing loss (SSD excluded); the subset used in each analysis is determined by data availability for that task. The subsets are not nested in all cases, and consistency of conclusions across overlapping samples was verified throughout.

| **Analysis** | **RQ** | $N$ | **Predictors** | **Outcome** | **Reported in** |
|---|---|---|---|---|---|
| Cohort description | — | 473 | — | — | Table 1 |
| Outcome variability | — | 378 | — | Post-op WRS | Results, Sec. 3.1 |
| Individual score prediction | RQ1 | 378 | 57 pre-op features (3, 7, 17, 57 subsets) | Post-op WRS | Table 2 |
| Validation hierarchy | RQ1 | 378 | 57 pre-op features | Post-op WRS | SI Sec. S4 |
| Learning curve (25–100% subsamples) | RQ1 | 94–378 | 57 pre-op features | Post-op WRS | Results, SI Sec. S4 |
| Imputation comparison (5 strategies) | RQ1 | 378 | 57 pre-op features | Post-op WRS | SI Sec. S8 |
| Paired sensitivity, pre-op WRS added | RQ1 | 264 | 57 features + pre-op WRS | Post-op WRS | Results, SI Sec. S4 |
| *Pre-operative control* | RQ1 | 294 | 57 pre-op features; audiometry only | *Pre*-op WRS | Results, Sec. 3.2 |
| Correlation, PTA vs speech scores | RQ1 | 262/366 | Pre-op PTA | Pre- and post-op WRS | Results, Sec. 3.3 |
| Distributional analysis ($W_1$) | RQ1 | 264 | 46 features, one at a time | Pre- and post-op WRS | Table S9, Fig. S3 |
| Risk classification | RQ2 | 378 | 57 pre-op features | Poor outcome (<40%) | Results, Sec. 3.4 |
| Univariate screen, Bonferroni $k = 49$ | RQ2 | varies | 49 features, one at a time | Poor outcome | Table S9 |
| AUC by feature subset | RQ2 | 378 | 35 / 6 / 5 / 46 | Poor outcome | Fig. 2, Table S10 |
| Threshold sensitivity (30–50%) | RQ2 | 378 | 57 pre-op features | Poor outcome | Table S5 |
| Calibration | RQ2 | 378 | 57 pre-op features | Poor outcome | Table S6 |
| Six-month prognostic value | RQ3 | 188 | 57 features ± six-month score | Long-term WRS | Results, Sec. 3.5 |
| *Six-month risk classification* | RQ3 | 188 | 57 features ± six-month score | Poor long-term outcome | Results, Sec. 3.5 |
| Trajectory clustering | RQ3 | 136 | Serial WRS scores | Trajectory type | Fig. 4 |
| Trajectory prediction | RQ3 | 136 | 57 pre-op features | Trajectory type | Results, Sec. 3.5 |
| Within-session reliability | — | 294/378 | WRS list 1 | WRS list 2 | Table S8 |
| Availability of the outcome | — | 398 | Pre-op profile | Whether tested | SI Sec. S9 |

Models: all prediction analyses use the seven algorithms of Table 2 unless stated; where a single value is quoted in the text it is the best-performing model, TabPFN. Italicised rows are analyses added in response to reviewer comments.

### SI Section S2: Distributional analysis of post-operative redistribution

This section reports the full distributional analysis summarised in Section 3.3 of the main text. Patients were stratified by each pre-operative variable and the optimal-transport (1-dimensional Wasserstein) distance $W_1$ between strata was computed separately for pre- and post-operative Mono1 distributions; the relative change $(W_{1,\text{pre}} - W_{1,\text{post}})/W_{1,\text{pre}}$ was tested by permutation (5,000 label permutations). Of the 46 features for which a distance could be computed, the between-group separation compresses after implantation for 23 (all audiometric, aided-field and pre-operative speech measures), amplifies for 7 (onset of deafness in either ear, sign language history, learned profession, education, multilingual home, family history of CI use), and shows no significant change for 16 (Supplementary Material, Figure S3 and Table S9). The compression reflects convergence of group means, not of individual spread: across the 23 compressing variables, mean within-stratum SD fell only from 27.4 to 25.0 percentage points (9%), while mean between-stratum $W_1$ fell from 21.5 to 6.0 (72%).

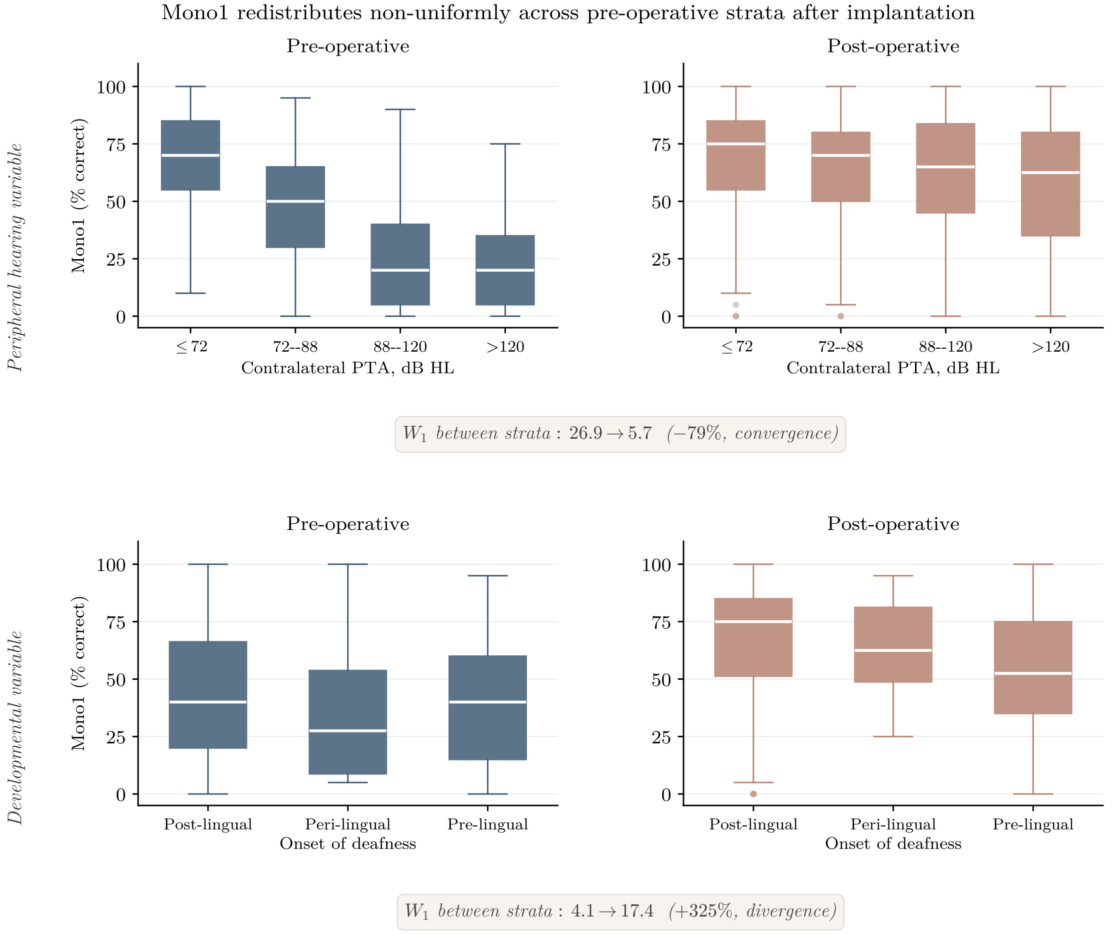


Figure S1: **Post-operative redistribution of Mono1 across pre-operative strata.** Boxplots of Mono1 (% correct) stratified by two representative pre-operative variables, before and after implantation. *Top row*: contralateral four-frequency PTA, in quartiles (≤72, 72–88, 88–120, >120 dB HL). Pre-operatively (left), Mono1 medians decline monotonically across quartiles from 70% to 20%, the standard audiometric-perceptual relationship. Post-operatively (right), the same quartiles compress to 75–62%: mean between-stratum $W_1$ falls from 26.9 to 5.7 (−79%, permutation $p < 0.001$). *Bottom row*: onset of deafness. Pre-operative distributions overlap closely across categories ($W_1 = 4.1$); post-operatively they separate, with post-lingual onset highest and pre-lingual lowest, and $W_1$ rising to 17.4 (+325%, $p < 0.001$). The two rows illustrate the selective redistribution summarised in Table S2: peripheral hearing variables lose between-stratum separation after implantation, developmental variables gain it.

### SI Section S3: TabPFN fine-tuning details

Fine-tuning was performed on an NVIDIA DGX Spark workstation (GB10 architecture, CUDA 13.0) with 50 epochs, learning rate $5 \times 10^{-6}$ (selected by grid search over $\{5\text{e-}6, 1\text{e-}5, 5\text{e-}5, 1\text{e-}4, 5\text{e-}4\}$), weight decay 0.01, AdamW optimizer, with 2 ensemble members during fine-tuning and validation and 4–8 at final inference.

Table S2: Illustrative variables from the Equalizer/Discriminator/Persistence framework, drawn from the 57-variable pre-operative pool. $W_{1,\text{pre}}$ and $W_{1,\text{post}}$ are mean pairwise Wasserstein distances between strata. Relative change $\Delta W_1(\%) = (W_{1,\text{pre}} - W_{1,\text{post}})/W_{1,\text{pre}} \times 100$; positive values indicate convergence. $p$-value from permutation testing (5,000 label permutations); for Equalizer variables, $p$ tests convergence; for Discriminator, divergence; for Persistence, neither direction was significant. The full variable-level results are available from the authors.

| Variable | Category | $W_{1,\text{pre}}$ | $W_{1,\text{post}}$ | $\Delta W_1$ (%) | $p$ |
|---|---|---|---|---|---|
| *Equalizer (between-group convergence; 24 of 57 variables)* | | | | | |
| Phone use | Communication | 14.8 | 2.0 | −86 | < 0.001 |
| Co PTA4 | Audiometric | 26.9 | 5.7 | −79 | < 0.001 |
| Bilateral CI | Clinical | 7.6 | 2.6 | −66 | 0.045 |
| CI PTA4 | Audiometric | 13.7 | 8.3 | −39 | 0.006 |
| *Discriminator (between-group divergence; 8 of 57 variables)* | | | | | |
| Onset of deafness | HL history | 4.1 | 17.4 | +325 | < 0.001 |
| Sign language | Communication | 9.7 | 20.5 | +112 | 0.006 |
| Profession learned | Socioeconomic | 6.8 | 13.1 | +94 | 0.003 |
| *Persistence (no significant change; 25 of 57 variables)* | | | | | |
| Articulation | Communication | 18.0 | 19.4 | −8 | 0.77 |
| Duration of HL | Demographic | 8.1 | 8.5 | −4 | 0.73 |
| CI PTA 250 Hz | Audiometric | 5.5 | 5.5 | 0 | 0.60 |

### SI Section S4: Robustness analyses

**The ceiling holds across validation strategies.** We further evaluated linear regression, Random Forest, and TabPFN on the 17-feature primary configuration across the validation hierarchy from apparent fit to temporal generalisation [Collins et al., 2024, Steyerberg, 2019, Ramspek et al., 2021]. For Random Forest, every honest generalisation estimate (a single 70/30 hold-out split averaged across 20 random seeds, 10-fold cross-validation, 5-fold cross-validation, leave-one-out cross-validation, and a temporal split training on patients implanted before 2022 and testing on those implanted from 2022 onward) returned $R^2$ at or below zero, with values clustered between $-0.04$ and $-0.06$ across the five honest validation strategies. The same pattern held for linear regression, with cross-validated $R^2$ between $-0.01$ and $-0.07$. The temporal split was the most pessimistic condition for both algorithms (linear $-0.07$, Random Forest $-0.21$). The pattern is therefore robust to the choice of validation strategy.

**Sensitivity analyses.** Several sensitivity checks confirm the limit. Adding more features did not improve performance: cross-validated $R^2$ worsened with feature-set size for the linear models. Lasso regularisation, which selects the most informative features automatically, returned $R^2 \approx +0.01$, no better than the unselected 7-feature version. Alternative outcome formulations did not help either: log-transformed Mono1, arcsine-square-root Mono1, and the change score (post − pre) all returned cross-validated $R^2$ at or below the absolute-Mono1 baseline. Including the pre-operative Mono1 score in the smaller paired-sample analysis ($N = 264$) yielded a TabPFN $R^2 \approx 0.10$, with the conventional algorithms returning $R^2$ at or below zero.

**Univariate odds-ratio robustness.** A further check concerned the univariate odds-ratio analysis rather than the regression. Excluding unmeasurable audiometric and aided-field thresholds, rather than imputing them, does not change which variable survives Bonferroni correction, whereas imputing them introduces a spurious survivor (Table S3).

Table S3: Sensitivity of the univariate poor-outcome analysis to the handling of unmeasurable audiometric and aided-field thresholds ($N = 378$; Bonferroni threshold $0.05/49 = 0.00102$). Only the audiometric and aided-field variables are affected; the developmental and communicative variables are unchanged. Under the primary analysis (unmeasurable excluded) and under the raw scores, *COM_ARTICULATION* is the only variable to survive Bonferroni correction; imputing unmeasurable thresholds to 130 or 115 dB HL causes *prFF_2000_CI* to additionally cross the threshold, an artefact of the imputed block of identical extreme values.

| **Handling of unmeasurable** | **audio/FF nominal** ($p < 0.05$) | **audio/FF survive Bonf.** | **prFF_2000_CI** $p$ | **Survive Bonferroni** |
|---|---|---|---|---|
| Raw (sentinels retained) | 6 | 0 | 0.0050 | COM_ARTICULATION |
| Imputed to 130 dB HL | 13 | 1 | 0.0008 | prFF_2000_CI, COM_ARTICULATION |
| Imputed to 115 dB HL | 13 | 1 | 0.0008 | prFF_2000_CI, COM_ARTICULATION |
| Unmeasurable excluded | 4 | 0 | 0.0299 | COM_ARTICULATION |

### SI Section S5: Sample size, threshold, calibration, imputation, and reliability

Table S4: Cross-validated $R^2$ for post-operative WRS as a function of training sample size (fine-tuned TabPFN; 5 random subsamples per size, 10-fold cross-validation within each; mean ± SD across draws).

| **Fraction of cohort** | $N$ | **CV $R^2$ (TabPFN)** |
|---|---|---|
| 25% | 94 | $-0.084 \pm 0.106$ |
| 50% | 189 | $+0.039 \pm 0.058$ |
| 75% | 283 | $+0.026 \pm 0.049$ |
| 100% | 378 | $+0.033 \pm 0.006$ |

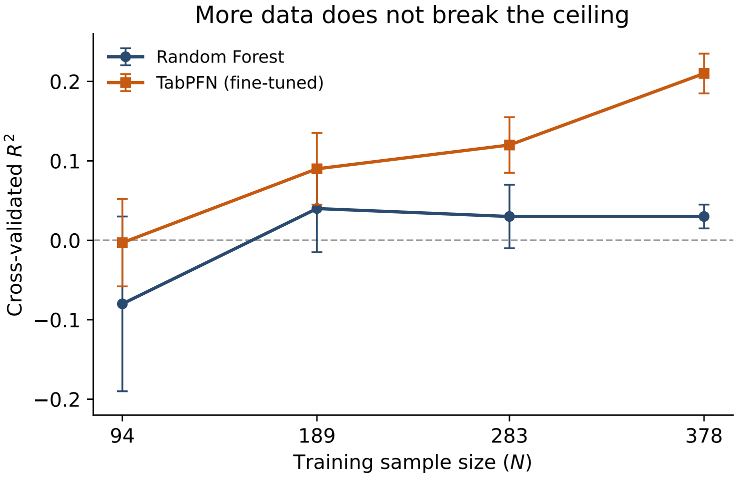


Figure S2: **Cross-validated $R^2$ as a function of training sample size.** Random Forest and fine-tuned TabPFN regression of post-operative WRS on the pre-operative feature set, evaluated on random subsamples of the cohort (5 random draws per size, 10-fold cross-validation within each). Error bars: standard deviation across draws. The Random Forest is flat from approximately $N = 190$ onward. The fine-tuned TabPFN continues to improve with sample size but reaches only $R^2 \approx 0.22$ at the full cohort, still leaving nearly four fifths of the variance in individual outcome unexplained. Over the sample sizes available here, added patients of the same kind do not lift either model to clinically useful accuracy; behaviour beyond the present cohort is untested.

Table S5: Cross-validated AUC for poor-outcome classification across alternative WRS thresholds ($N = 378$; 10-fold stratified cross-validation; balanced class weights). Discrimination is modest and broadly stable across thresholds; the 40% cutoff used in the main analysis is not a favourable special case.

| **Threshold** | ***N* poor** | **% poor** | **AUC (LR)** | **AUC (RF)** |
|---|---|---|---|---|
| $< 30\%$ | 53 | 14.0% | 0.594 | 0.659 |
| $< 35\%$ | 63 | 16.7% | 0.592 | 0.555 |
| $< 40\%$ | 75 | 19.8% | 0.651 | 0.587 |
| $< 45\%$ | 87 | 23.0% | 0.592 | 0.576 |
| $< 50\%$ | 106 | 28.0% | 0.588 | 0.575 |

Table S6: Calibration of the poor-outcome classifiers (WRS $< 40\%$, $N = 378$, 10-fold cross-validated out-of-fold probabilities). Predicted versus observed risk by quartile of predicted probability. The logistic model with balanced class weights over-states risk substantially in the upper quartiles, and its Brier score is worse than that of a predictor that always returns the cohort prevalence (0.159).

| **Model** | **AUC** | **Brier** | **Quartile** | **Predicted / Observed** |
|---|---|---|---|---|
| Logistic regression | 0.667 | 0.222 | Q1 | 0.10 / 0.09 |
| | | | Q2 | 0.25 / 0.18 |
| | | | Q3 | 0.46 / 0.19 |
| | | | Q4 | 0.79 / 0.33 |
| Random Forest | 0.587 | 0.156 | Q1 | 0.08 / 0.13 |
| | | | Q2 | 0.15 / 0.24 |
| | | | Q3 | 0.21 / 0.15 |
| | | | Q4 | 0.35 / 0.27 |
| Prevalence-only baseline | — | 0.159 | — | 0.20 / 0.20 |

Table S7: Performance across imputation strategies for the pre-operative predictors ($N = 378$; Random Forest regression of WRS and logistic-regression classification of poor outcome, both 10-fold cross-validated). All strategies return cross-validated $R^2$ near zero and comparable AUC, confirming that the limit is not an artefact of the imputation choice.

| **Imputation** | **CV $R^2$ (regression)** | **CV AUC (classification)** |
|---|---|---|
| Mean | +0.054 | 0.659 |
| Median (primary) | +0.004 | 0.667 |
| $k$-NN ($k = 5$) | +0.008 | 0.639 |
| Iterative (MICE) | +0.064 | 0.640 |

Table S8: Within-session reliability of the Freiburger monosyllable test, estimated from the two equivalent 20-word lists administered in the same session. Reliability is essentially identical before and after implantation, so the change in prognostic value of the test after surgery, described in "Changes to Predictor-Outcome Relationship" in the main text, cannot be attributed to a change in measurement precision.

| **Regime** | ***N*** | **Pearson *r*** | **ICC(2,1)** |
|---|---|---|---|
| Pre-operative (best-aided) | 294 | 0.922 | 0.922 |
| Post-operative (CI only) | 378 | 0.921 | 0.921 |

**SI Section S6: Full predictor importance**

Table S9: Full predictor importance for poor outcome classification (WRS $< 40\%$, $N = 378$, 75 poor outcomes) and change in between-group separation after implantation. Univariate logistic regression odds ratios with 95% CI and $p$-values. For audiometric and aided-field variables, unmeasurable thresholds were excluded from the univariate estimation (measurable thresholds only), so the odds ratio is per decibel over the measurable range and $N$ varies accordingly. $W_1$ is the size-weighted mean optimal-transport distance between the WRS distributions of the groups a feature defines, in percentage points of the score, computed on the paired subsample ($N = 264$) before and after implantation; continuous features are split at quartiles and groups smaller than 10 patients are not used. $p_{\text{perm}}$ is from 5,000 label permutations and tests the change from $W_{1,\text{pre}}$ to $W_{1,\text{post}}$ in whichever direction it occurred. Multivariate ranks: absolute standardized LR coefficient, RF permutation importance, and TabPFN permutation importance. Variables sorted by univariate $p$-value. Bonferroni threshold ($k = 49$, the number of pre-operative variables for which a univariate model could be fitted): $p < 0.00102$.

| | | | | Between-group separation | | | Multivariate rank | | |
|---|---|---|---|---|---|---|---|---|---|
| **Variable** | $N$ | **OR (95% CI)** | $p$ | $W_{1,\text{pre}}$ | $W_{1,\text{post}}$ | $p_{\text{perm}}$ | **LR** | **RF** | **TabPFN** |
| **COM_ARTICULATION** | **368** | **2.55 [1.70, 3.81]** | **<0.001** | **18.3** | **19.7** | **0.210** | **1** | **2** | **1** |
| SES_PROFESSION_LEARNED | 325 | 0.73 [0.61, 0.88] | 0.0012 | 6.8 | 11.5 | 0.003 | 16 | 10 | 9 |
| prPTA_1000_CI | 313 | 1.03 [1.01, 1.05] | 0.005 | 12.4 | 8.1 | 0.017 | 37 | 21 | 10 |
| COM_Gebaerden | 355 | 3.36 [1.42, 7.94] | 0.006 | 9.3 | 20.2 | 0.007 | 20 | 33 | — |
| prPTA_2000_CI | 248 | 1.04 [1.01, 1.06] | 0.006 | 12.9 | 7.0 | 0.004 | 11 | 10 | 3 |
| SES_EDUCATION_LEVEL | 346 | 0.76 [0.62, 0.93] | 0.009 | 7.0 | 10.6 | 0.014 | 36 | 3 | — |
| COM_MULTILANG_HOME | 367 | 1.74 [1.15, 2.64] | 0.009 | 4.2 | 11.0 | 0.008 | 19 | 21 | 5 |
| prPTA_6000_Co | 213 | 1.03 [1.00, 1.05] | 0.021 | 23.2 | 7.3 | <0.001 | 2 | 33 | — |
| EVA_DEAF_ONSET | 332 | 1.99 [1.09, 3.64] | 0.025 | 3.6 | 17.1 | <0.001 | 21 | 41 | — |
| prFF_2000_CI | 205 | 1.02 [1.00, 1.04] | 0.030 | 7.0 | 7.2 | 0.373 | 30 | 41 | 7 |
| EVA_DEAF_ONSET_L | 285 | 2.06 [1.06, 4.01] | 0.033 | 6.7 | 19.0 | <0.001 | 15 | 21 | — |
| Mono2_pre | 264 | 0.99 [0.97, 1.00] | 0.037 | 37.2 | 7.2 | <0.001 | 26 | 10 | 12 |
| COM_FATHER_HACI_USER | 139 | 0.27 [0.08, 0.97] | 0.045 | 4.8 | 15.6 | 0.003 | 6 | 33 | 8 |
| prPTA_500_CI | 341 | 1.01 [1.00, 1.03] | 0.053 | 7.8 | 5.7 | 0.171 | 8 | 10 | — |
| prPTA_1000_Co | 336 | 1.01 [1.00, 1.03] | 0.065 | 23.2 | 5.1 | <0.001 | 13 | 21 | — |
| C12_pre | 255 | 0.99 [0.97, 1.00] | 0.081 | 31.4 | 6.2 | <0.001 | 5 | 6 | — |
| prPTA_4000_CI | 193 | 1.03 [1.00, 1.06] | 0.090 | 8.7 | 5.6 | 0.075 | 32 | 3 | — |
| prFF_6000_Co | 188 | 1.01 [1.00, 1.03] | 0.091 | 17.1 | 5.7 | <0.001 | 24 | 10 | — |
| prFF_500_Co | 264 | 0.98 [0.96, 1.00] | 0.095 | 11.4 | 7.0 | 0.014 | 12 | 21 | — |
| prFF_1000_CI | 239 | 1.02 [1.00, 1.04] | 0.116 | 6.2 | 3.9 | 0.147 | 39 | 33 | — |
| FM_pre | 274 | 0.98 [0.95, 1.01] | 0.125 | 21.1 | 2.4 | <0.001 | 35 | 21 | 14 |
| prPTA_500_Co | 343 | 1.01 [1.00, 1.02] | 0.147 | 17.9 | 6.3 | <0.001 | 14 | 41 | — |
| EVA_HL_PROGREDIENT | 337 | 0.41 [0.12, 1.38] | 0.147 | 7.6 | 4.0 | 0.193 | 17 | 6 | — |
| Mono1_pre | 264 | 0.99 [0.98, 1.00] | 0.153 | 40.6 | 6.6 | <0.001 | 40 | 48 | — |
| prPTA_2000_Co | 296 | 1.01 [1.00, 1.03] | 0.161 | 23.7 | 5.9 | <0.001 | 31 | 10 | — |
| prFF_4000_CI | 164 | 1.01 [0.99, 1.03] | 0.180 | 6.4 | 6.3 | 0.466 | 27 | 21 | — |
| prPTA_8000_Co | 102 | 1.03 [0.99, 1.07] | 0.187 | 26.8 | 7.8 | <0.001 | 29 | 6 | 15 |
| prFF_6000_CI | 123 | 1.02 [0.99, 1.04] | 0.224 | 4.8 | 5.6 | 0.284 | 34 | 10 | — |
| prPTA_4000_Co | 254 | 1.01 [0.99, 1.03] | 0.226 | 24.2 | 5.8 | <0.001 | 4 | 21 | — |
| Num_pre | 272 | 0.99 [0.98, 1.01] | 0.230 | 33.9 | 3.3 | <0.001 | 41 | 10 | — |
| prFF_250_Co | 259 | 0.99 [0.97, 1.01] | 0.235 | 8.5 | 5.6 | 0.088 | 45 | 41 | — |
| prFF_500_CI | 240 | 1.01 [0.99, 1.03] | 0.241 | 4.3 | 4.7 | 0.340 | 42 | 21 | — |
| prPTA_6000_CI | 131 | 1.02 [0.98, 1.06] | 0.298 | 8.8 | 5.2 | 0.089 | 44 | 10 | — |
| prFF_4000_Co | 218 | 1.01 [0.99, 1.02] | 0.314 | 21.4 | 7.6 | <0.001 | 18 | 6 | — |
| prFF_250_CI | 239 | 1.01 [0.99, 1.02] | 0.428 | 5.9 | 6.5 | 0.294 | 49 | 41 | — |
| prPTA_250_CI | 282 | 1.01 [0.99, 1.02] | 0.458 | 5.2 | 5.9 | 0.248 | 3 | 10 | — |
| V08_pre | 265 | 0.99 [0.98, 1.01] | 0.496 | 29.6 | 5.7 | <0.001 | 7 | 41 | — |
| prPTA_8000_CI | 52 | 1.03 [0.95, 1.11] | 0.515 | — | — | — | 38 | 41 | — |
| COM_PHONE_USE | 343 | 0.92 [0.70, 1.22] | 0.574 | 14.7 | 5.4 | <0.001 | 43 | 10 | — |
| prPTA_125_Co | 279 | 1.00 [0.99, 1.02] | 0.705 | 11.5 | 4.7 | <0.001 | 25 | 33 | 6 |
| prFF_1000_Co | 261 | 1.00 [0.97, 1.02] | 0.713 | 17.3 | 6.0 | <0.001 | 33 | 21 | — |
| prPTA_125_CI | 207 | 1.00 [0.98, 1.02] | 0.725 | 4.9 | 6.4 | 0.151 | 28 | 33 | — |
| COM_LIP_READING | 265 | 1.09 [0.53, 2.23] | 0.807 | 9.9 | 4.4 | 0.047 | 48 | 21 | — |
| Geschlecht | 378 | 1.06 [0.64, 1.76] | 0.819 | 1.7 | 4.1 | 0.095 | 46 | 5 | — |
| prPTA_250_Co | 321 | 1.00 [0.99, 1.01] | 0.846 | 13.9 | 4.5 | <0.001 | 9 | 33 | 11 |
| prFF_8000_Co | 19 | 0.99 [0.94, 1.05] | 0.864 | — | — | — | 47 | 49 | 4 |
| prFF_8000_CI | 12 | 0.99 [0.90, 1.10] | 0.897 | — | — | — | 22 | 33 | 13 |
| prFF_2000_Co | 249 | 1.00 [0.98, 1.02] | 0.908 | 19.9 | 7.0 | <0.001 | 23 | 21 | — |
| Age_at_OP | 378 | 1.00 [0.98, 1.01] | 0.913 | 8.6 | 5.3 | 0.064 | 10 | 1 | 2 |

*Bold row: Bonferroni-corrected significance ($p < 0.05/49 = 0.00102$); only COM_ARTICULATION meets it. LR and RF ranks from a single fit on the full cohort with median imputation of missing values (RF: 500 trees, balanced class weights, permutation importance, 30 repeats). TabPFN ranks: top 15 from the fine-tuned TabPFN run, '—' below, carried pending a refreshed run. Only COM_ARTICULATION survives Bonferroni correction; the AUC of the multivariate models (TabPFN $= 0.71$) thus reflects non-linear interactions across features rather than additional independent main-effect predictors.*

A dash in the $W_1$ columns marks features for which the groups were too small to compute a distance (prPTA_8000_CI, prFF_8000_Co, prFF_8000_CI). Of the 46 features with a distance, 23 converge after implantation, 7 move apart and 16 show no significant change;

the pattern is shown in Supplementary Material, Figure S3. The feature subsets used for classification comprise 46 of the 49 features listed here: two features could not be assigned to a subset, as well as pre-operative WRS. Pre-operative WRS is listed here because each feature is tested separately, which is possible on the subsample with both a pre- and a post-operative score ($N = 264$; note the reduced $N$ in its row). It is not used as a predictor in the multivariable models or in the trajectory analyses, where requiring paired scores would reduce the cohort from 378 to 264.

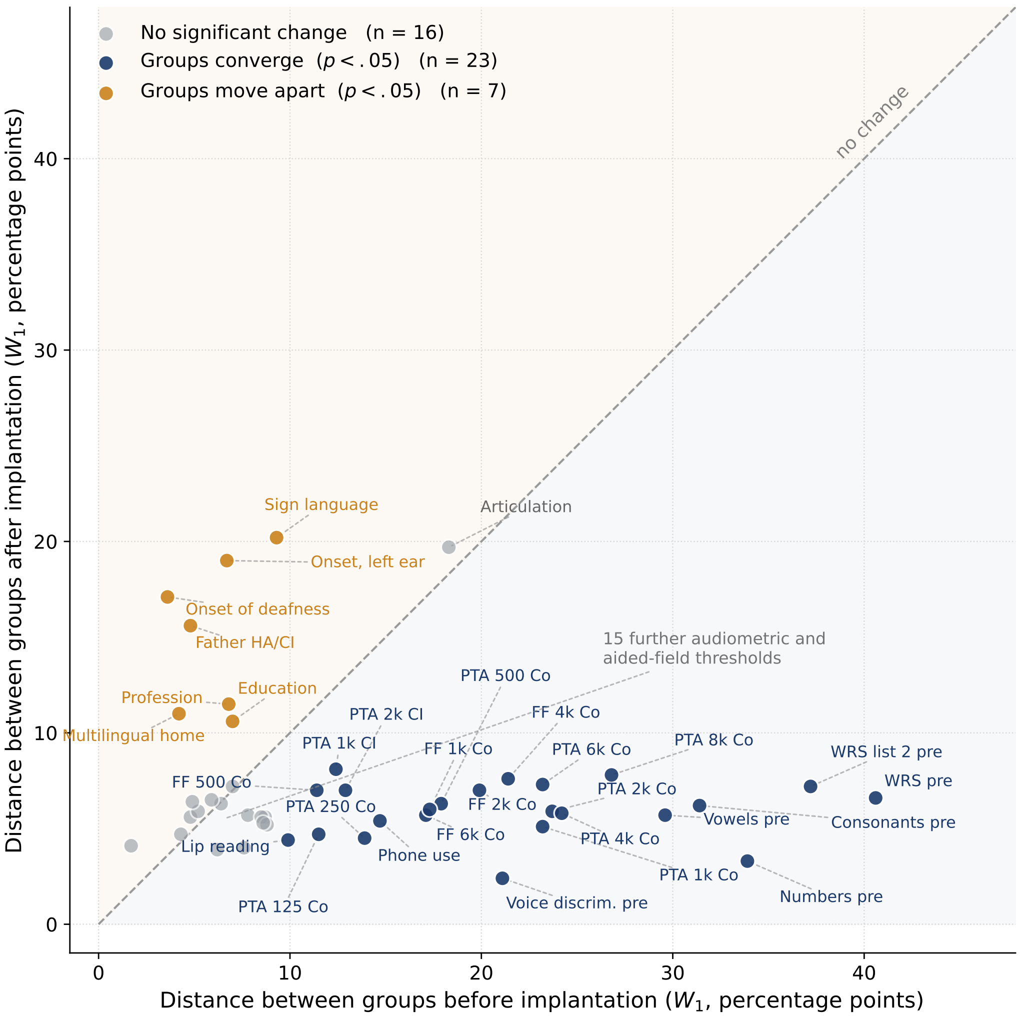


Figure S3: Change in between-group separation after implantation, for the 46 pre-operative features of Table S9 for which a distance could be computed. Each point is one feature. The horizontal axis is the size-weighted mean optimal-transport distance ($W_1$) between the WRS distributions of the groups that feature defines, before implantation; the vertical axis is the same distance after implantation, both in percentage points of the score. The dashed diagonal marks no change: points below it belong to features whose groups converge after implantation, points above to features whose groups move apart. Colour indicates the permutation test (5,000 label permutations, $p < .05$). Twenty-three features converge, seven move apart and sixteen show no significant change. The features that converge are the audiometric, aided-field and pre-operative speech measures; those that move apart are developmental and communicative. Articulation is the exception, unchanged and with the second largest post-operative separation in the pool. Fifteen further audiometric and aided-field thresholds sit in the low-low corner and are not labelled individually; their values are in Table S9.

### SI Section S7: Classification performance by pre-operative variable type

This table was presented in the main text of earlier versions and is reproduced here in full, with fold-level variability, to meet the journal's limit of three tables in the manuscript. Figure 2 of the main text displays the same AUC values graphically.

Table S10: Cross-validated AUC for poor absolute outcome (Mono1 $< 40\%$) by feature subset and algorithm. $N = 378$ adults, 75 (19.8%) poor outcomes; 10-fold stratified cross-validation. Subsets grouped by clinical variable type. Audiometric-only logistic regression is essentially at chance (AUC $= 0.53$), in contrast with the developmental and communicative subsets.

| Feature subset | # vars | LR | Random Forest | TabPFN |
|---|---|---|---|---|
| Audiometric, aided field, pre-op speech | 35 | $0.529 \pm 0.08$ | $0.587 \pm 0.06$ | $0.585 \pm 0.09$ |
| Developmental and communicative | 6 | $0.641 \pm 0.13$ | $0.577 \pm 0.09$ | $0.654 \pm 0.13$ |
| Demographic and clinical | 5 | $0.650 \pm 0.15$ | $0.594 \pm 0.12$ | $0.666 \pm 0.16$ |
| **All combined** | 46 | $0.667 \pm 0.13$ | $0.613 \pm 0.08$ | $0.657 \pm 0.14$ |

*Values: mean ± SD across 10 folds. All models, TabPFN included, were fine-tuned on the present dataset; the full-set TabPFN classifier reached AUC* $= 0.71$ *(reported in the text). All three algorithms reproduce the same ordering, with audiometric predictors at chance and developmental or communicative predictors above chance.*

### SI Section S8: Extended methods

This section holds material moved from the Methods of the main text to meet the journal's word limit. No content has been changed.

**Speech tests.** Mono1 (Freiburger monosyllables) is an open-set test in which the patient repeats monosyllabic German nouns and the tester records the response [Hahlbrock, 1953]; two equivalent lists of 20 words each are presented in the same session; Mono1 denotes the score on the first list and Mono2 the score on the second, each expressed as the percentage of words correctly identified (word recognition score). Mono2 is used in this study to estimate the within-session reliability of the test (Table S8) and appears among the pre-operative predictors. Mono1 is a mandatory outcome measure across all five Swiss CI centres [Brand et al., 2014] and serves as the primary outcome in the present study. V08 is a closed-set vowel identification task in which the patient selects one of eight vowels presented as CV logatomes in a /d_/ frame (e.g., /da/, /di/, /do/); it is primarily sensitive to spectral resolution, specifically F1/F2 formant contrasts [Dorman et al., 1991]. C12 is a closed-set consonant identification task in which the patient selects one of 12 consonants presented as VCV logatomes in an /a_a/ frame (e.g., /aba/, /ada/, /aga/); it is primarily sensitive to place-of-articulation contrasts (F2 transitions) [Rødvik et al., 2018]. FM (FEMMAL voice discrimination) is a yes/no task in which the patient identifies whether a speaker is male or female across 20 sentences; it is sensitive to fundamental frequency and temporal cues [Fu et al., 2005]. Num is an open-set task in which the patient repeats 20 two-digit numbers. V08, C12, FM, and Num are part of the German Minimal Auditory Capability (MAC) test battery [Dillier and Spillmann, 1992].

Post-operative Mono1 was available for 236 adults at 0–6 months, 267 at 6–12 months, 317 at 12–24 months, and 273 beyond 24 months (SSD excluded). These counts are not mutually exclusive: patients with repeated follow-up contribute to more than one interval.

**The 57 pre-operative features.** These comprised: pure-tone audiometric thresholds at individual frequencies (125–8000 Hz) for both the implanted and contralateral ear, aided free-field thresholds (250–6000 Hz), four-frequency PTA for both ears, pre-operative scores on the four speech tests other than WRS, age at implantation, sex, onset of deafness (post-lingual, peri-lingual, pre-lingual), duration of hearing loss, etiology, articulation quality (normal, slightly disturbed, strongly disturbed), progressive hearing loss (yes/no), CT/MRI findings, and socioeconomic and communication variables (education, living situation, insurance, phone use, lip reading, native language). Feature sets of varying size were constructed from this pool to test sensitivity to feature selection; the prediction methodology, imputation strategy, and modelling algorithms are described in Section 2.2 of the main text. The final feature set and its justification are presented in the Results. Of these 57 features, 49 could be fitted in a univariate logistic model of poor outcome and constitute the set over which the Bonferroni correction in the risk analysis is computed ($k = 49$;

Section 3.3 of the main text); the remaining 8 do not yield a single univariate odds ratio (nominal categorical variables with several unordered levels, such as etiology and native language, or variables with a degenerate near-constant distribution) and are therefore absent from the univariate column of Table S9.

**Imputation strategies, algorithms and validation.** Feature sets ranged from 3 clinical variables to all 57 pre-operative features with pairwise interaction terms, including a 7-feature clinical set and data-driven selection via adaptive best-subset selection (ABESS) and Lasso regularisation. Missingness in pre-operative predictors ranged from 25% to 40%; post-operative outcomes were never imputed. Five imputation strategies were compared (mean, median, $k$-nearest neighbours, multiple imputation by chained equations, and iterative imputation); the primary analyses use median imputation. Eight algorithms spanning three model families were applied to each feature-imputation combination: penalised linear models (linear, ridge, and lasso regression), the models most commonly reported in earlier CI outcome studies [Lazard et al., 2012, Blamey et al., 2013]; tree-based ensembles (random forests, gradient boosting, XGBoost, LightGBM), which capture non-linear interactions and have been applied to CI outcome prediction more recently [Shafieibavani et al., 2021, Demyanchuk et al., 2025]; and TabPFN [Hollmann et al., 2025], a foundation model that is among the top-performing methods for small-to-medium tabular data and the most flexible model in our grid. We evaluated increasingly flexible models, culminating in TabPFN, to test whether greater modelling capacity could overcome the ceiling, not to advocate any single method. TabPFN was fine-tuned on the present dataset for both the regression and the binary classification tasks, with cross-validation folds fixed before fine-tuning so that test-fold patients were never seen during training (Section S3). This grid ensures that any conclusion about predictability is not contingent on a particular modelling choice.

All results are reported under $k$-fold cross-validation ($k = 10$; $k = 5$ for the smaller 264-patient sample), avoiding the upward bias of in-sample $R^2$ that affects most published CI prediction models [van de Velde et al., 2021, Steyerberg, 2019]. To probe robustness to validation methodology, we also applied a 70/30 hold-out split (mean over 20 random splits), 5-fold and leave-one-out cross-validation, and a temporal split training on patients implanted before 2022 and testing on those implanted from 2022 onward; these span the validation hierarchy from apparent performance to external validation [Collins et al., 2024, Steyerberg, 2019, Ramspek et al., 2021]. We also computed 90% prediction intervals (via conformal prediction) giving, for each patient, the range expected to contain their true score 90% of the time.

#### SI Section S9: Availability of the post-operative outcome

Post-operative Mono1 was missing for 95 of the 473 adults in the study cohort. In total, 66 adults were implanted after June 2024 and had not yet reached a scheduled follow-up measurement at data extraction; 33 of these had no post-operative Mono1 score. Among the remaining 407 adults, implanted earlier, availability of the outcome was not independent of the pre-operative profile: adults without a post-operative Mono1 score had higher pre-operative PTA in the implanted ear (median 113.1 versus 102.5 dB HL, Mann-Whitney $p < .001$), were less often native speakers of German or Swiss German (67.1% of non-German speakers were tested versus 88.4% of German speakers, $p < .001$), and more often had disturbed articulation (74.4% and 75.0% tested for slightly and strongly disturbed articulation versus 88.7% for normal, $p = .002$). In a logistic model of testing status ($N = 398$) the three were independently associated (per 10 dB of pre-operative PTA, OR = 0.80, 95% CI 0.66 to 0.96; German or Swiss German main language, OR = 3.79, 2.02 to 7.09; per level of articulation disturbance, OR = 0.59, 0.38 to 0.93), while age at implantation was not.
